# Multi-ancestry genome-wide association study and meta-analysis of stimulant use disorder reveals biology and relationships to other psychiatric disorders

**DOI:** 10.64898/2026.06.05.26354997

**Authors:** Sarah E. Beck, Joseph D. Deak, Daniel F. Levey, Tian Ge, Paul W. Jeffries, Dongbing Lai, Travis T. Mallard, Louisa Degenhardt, Penelope A. Lind, Trine Tollerup Nielsen, Justin D. Tubbs, Leah Wetherill, Emma C. Johnson, Alexander S. Hatoum, The SUD Working Group of the Psychiatric Genomics Consortium, COGA Collaborators, Yale-Penn Collaboration, The VA Million Veteran Program, Anders D. Børglum, Ditte Demontis, Sarah E. Medland, Nicholas G. Martin, Elliot C. Nelson, Jordan W. Smoller, Henry R. Kranzler, J. Michael Gaziano, Murray B. Stein, Arpana Agrawal, Howard J. Edenberg, Joel Gelernter

## Abstract

Stimulant use disorder (StimUD) is a significant public health problem, but genetic studies have been limited by small sample sizes. We conducted genome-wide association studies (GWAS) of StimUD in the Million Veteran Program (MVP) and All of Us (AOU), followed by meta-analysis with FinnGen and 10 additional datasets, for a total of 709,369 individuals (N_cases_=33,977, N_controls_=675,392) in four broad ancestry groups: European (EUR) (N_cases_=22,564, N_controls_=624,672), African (AFR) (N_cases_=7,574, N_controls_=34,189), Admixed American (AMR) (N_cases_=3,657, N_controls_=15,698), and East Asian (EAS) (N_cases_=182, N_controls_=833). Population-specific SNP heritability was 6.1% in EUR and 2.4% in AFR. We discovered a total of 19 genome-wide-significant loci, six in EUR, including *DRD2*\*rs5794864, *P*=7.32×10^−10^, one in AFR, five in a multi-ancestry meta-analysis, including *CHRNA5*\*rs55781567, *P*=3.27×10^−9^, two in a male-only meta-analysis, including *FTO*\*rs8057044, *P*=9.50×10^−9^, and five in a meta-analysis of sex-stratified results. In a hold-out AOU subsample (N_EUR_=18,841, N_AFR_=12,263, N_AMR_=9,739), ancestry-specific polygenic risk scores were significantly associated with StimUD in EUR (OR=3.28, 95% confidence interval (CI)=2.89-3.71) and AMR (OR=2.01, 95% CI=1.71-2.37). Transcriptome-wide association studies, fine-mapping, and colocalization analyses prioritized additional genes (e.g., *GPX1*, *BSN*). Genetic correlation, Mendelian randomization, and causal mixture analyses revealed relationships with other substance use and use disorder phenotypes, including cannabis use disorder (r_g_=0.94, *P*=5.43×10^−237^) and opioid use disorder (r_g_=1.01, *P*=4.40×10^−107^), and other psychiatric traits, including anxiety, depression, neuroticism, and attention-deficit/hyperactivity disorder. This is the first well-powered GWAS of StimUD, and it offers significant insights into disease biology.

## Introduction

Stimulant use disorder (StimUD), including methamphetamine and prescription stimulant misuse, is a significant public health problem. (Cocaine use disorder (CocUD) is generally considered a separate diagnosis.) Overdose deaths in the U.S. involving stimulants alone and used with synthetic opioids rose from 5,716 in 2015 to 34,855 in 2023.^1^ The National Survey on Drug Use and Health (NSDUH) reports that, from 2021-2024, 2.4 million people, or 0.8% of the U.S. population aged 12 or older, used methamphetamine in the past year and 3.9 million people, or 1.4% of the population aged 12 or older, misused prescription stimulants.^2^ The prevalence of StimUD, a major complication of stimulant use, varies by sex and ancestry.^3^ NSDUH estimates that between 2015 and 2019, 0.4% of the European-ancestry (EUR) and Hispanic populations were diagnosed with methamphetamine use disorder in the past year, compared to 0.1% of the non-Hispanic African American and Asian populations, with similar rates for prescription stimulant use disorder (0.2% in whites, 0.1% in others).^3^ Among Medicaid enrollees aged 18-24, non-cocaine psychostimulant use disorder rates increased from 0.09% in 2000 to 0.49% in 2020.^4^

Twin studies demonstrate that StimUD is heritable (twin h^2^=42%)^5^, but insight into the genetics of stimulant use traits is limited. Prior genome-wide association studies (GWAS) of StimUD identified only one genome-wide significant (GWS) locus, *SLC25A16*\*rs2394476, *P*=3.42×10^−10^, in individuals of African ancestry (AFR), likely because of small sample size (total N_AFR_=5,758, total N_EUR_=5,681, total N in East Asians (EAS)=580).^6,7^ Previous work also identified risk loci for lifetime methamphetamine use (rs141493660, *P*=8.58×10^−10^, in admixed Americans (AMR) (N_eff_=8,007) and *EXT1*\*rs17431748, *P*=2.91×10^−8^, in a multi-ancestry meta-analysis (N_eff_=51,809)) and lifetime prescription stimulant use (*TBCD*\*rs55775765, *P*=4.45×10^−8^ in AMR (N_eff_=8,426) and *MEF2C-AS2*\*rs116758901, *P*=3.98×10^−8^ in a multi-ancestry meta-analysis (N_eff_=64,961)).^8^

To better understand the genetics and biology of StimUD, we performed GWAS and post-GWAS analyses in a meta-analyzed sample with a case count more than an order of magnitude larger than in previous studies: 33,977 cases and 675,392 controls (N_total_=709,369, N_eff_=108,543). Participants came from the Million Veteran Program (MVP)^9^, All of Us (AOU)^10^, FinnGen^11^, and 10 smaller datasets, including meta-analyses of EUR (N_case_=22,564, N_control_=624,672), AFR (N_case_=7,574, N_control_=34,189), and AMR (N_case_=3,657, N_control_=15,698) individuals, and a GWAS of EAS individuals in MVP (N_case_=182, N_control_=833). We also performed sex-specific GWAS. These analyses prioritized genes for further research and elucidated StimUD’s relationship to other psychiatric disorders.

## Results

### GWAS

647,236 EUR participants from 13 datasets were included in an effective-sample-size-weighted GWAS meta-analysis of StimUD (Table 1). Six independent (r^2^<0.1) GWS loci were identified in EUR (Table 2, Supplemental Figure 1). In the AFR meta-analysis of 41,763 participants from four datasets, one GWS locus was found (Table 2, Supplemental Figure 2). No GWS loci were identified in a meta-analysis of 19,355 AMR individuals in MVP and AOU or in a GWAS of 1,015 EAS individuals in MVP. Nine loci were significant in the multi-ancestry analysis, with five significant only in that analysis (Table 2, Supplemental Figure 3).

**Table 1:**
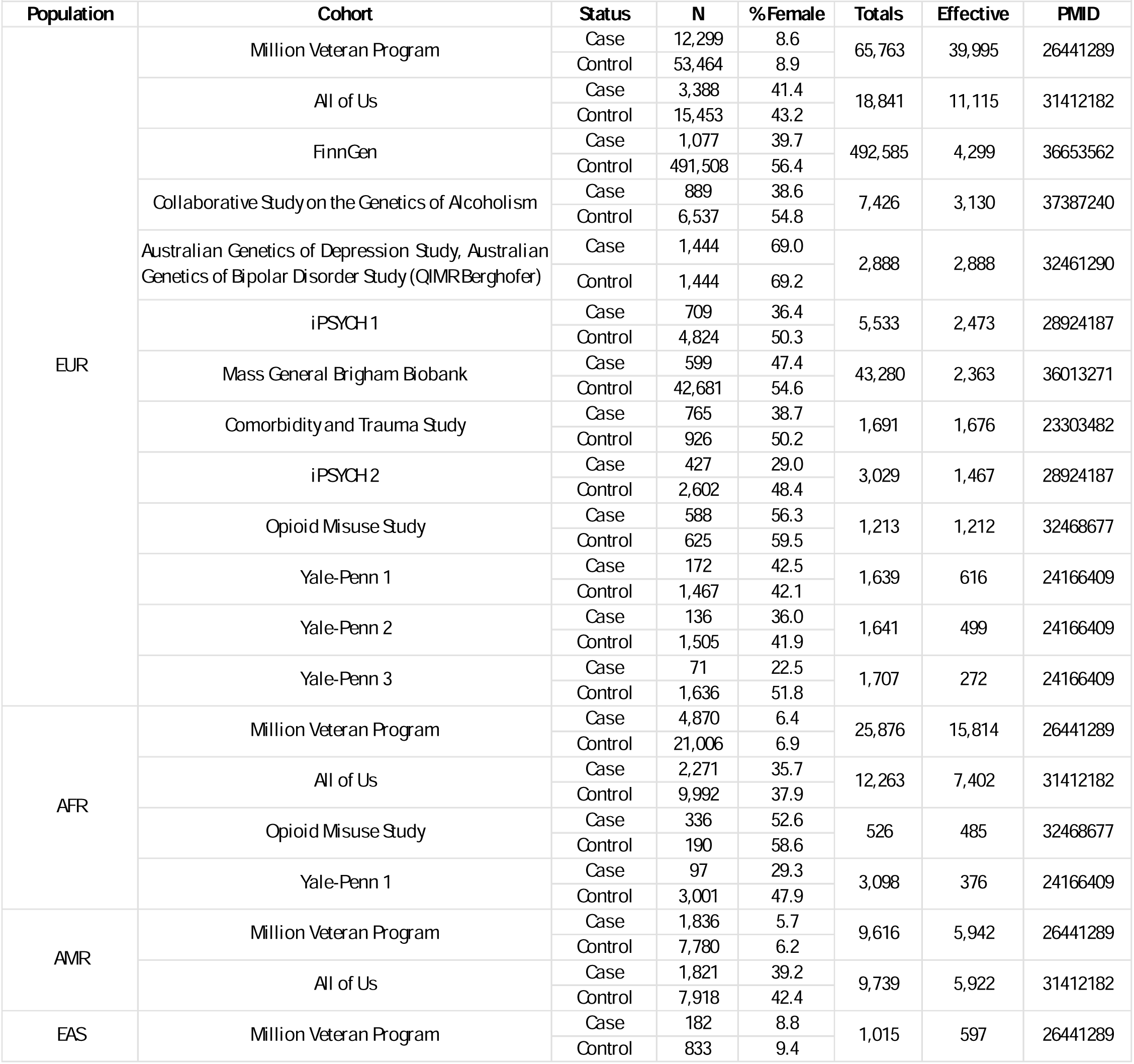
Demographics.

**Table 2:**
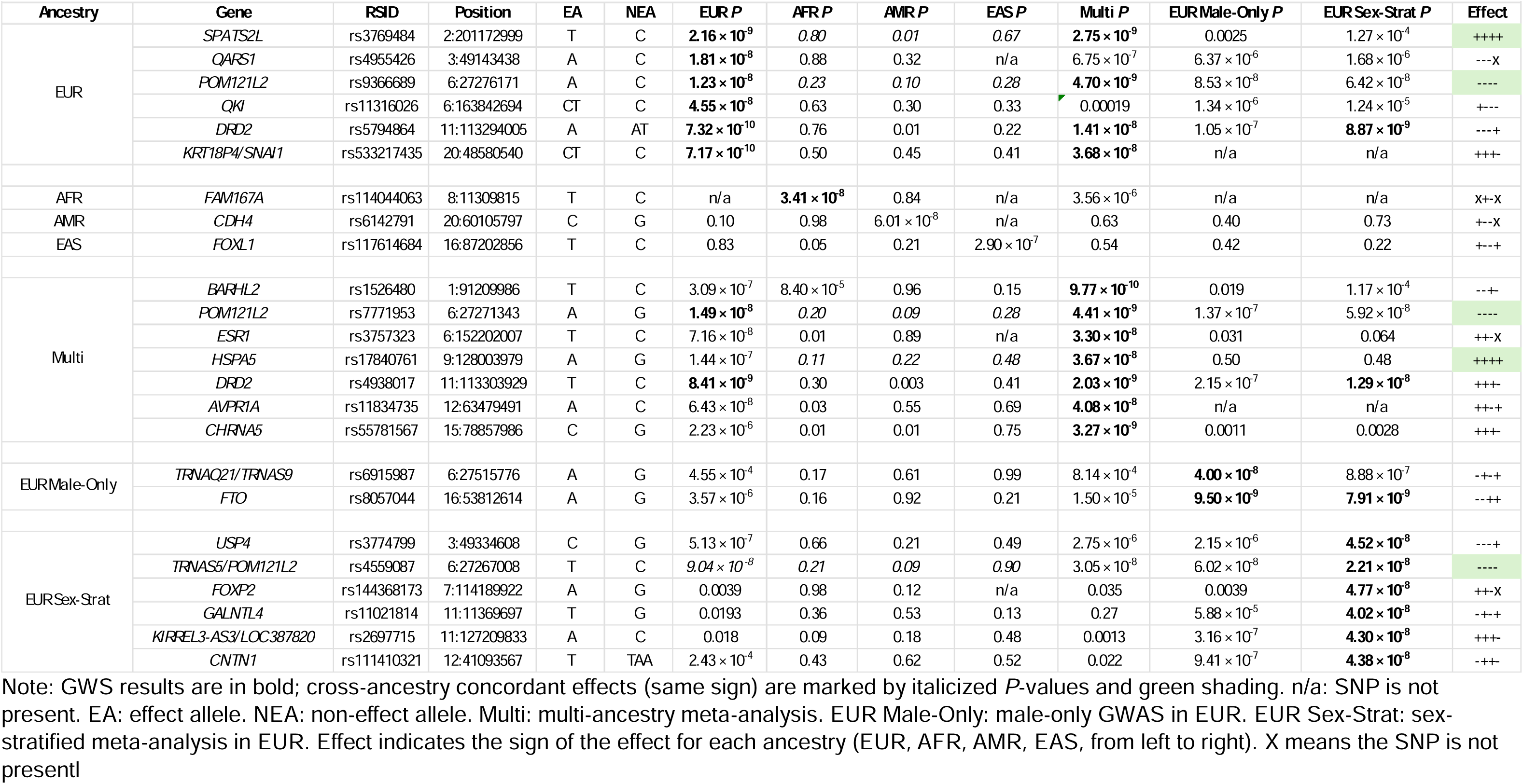
Lead SNPs for each ancestral group, the multi-ancestry meta-analysis, the male-only GWAS in EUR, and the sex-stratified meta-analysis in EUR.

One SNP identified in the EUR meta-analysis was nominally replicated (*P*<0.01) in AMR: *SPATS2L*\*rs3769484 (EUR β=0.032*, P=*2.16×10^−9^, AMR β=0.077*, P=*0.0069). *FAM167A*\*rs114044063, the SNP GWS in AFR, was present in AMR but not significant, possibly because of low sample size and minor allele frequency, and not present in EUR or EAS.

Of the SNPs GWS in the EUR and multi-ancestry meta-analyses, four (three independent) showed concordant effect direction across all ancestries: *SPATS2L*\*rs3769484, *HSPA5*\*rs17840761, and *POM121L2*\*rs9366689 and *rs7771953, which last are in close linkage disequilibrium (LD) (r^2^=0.99).

We then performed sex-stratified GWAS in EUR in MVP and AOU, the only datasets that had sufficient sample size to stratify by sex, and meta-analyzed the results. Among males (N_case_=13,206, N_control_=57,183), there were two GWS loci (Table 2, Supplemental Figure 4).

Among females (N_case_=2,454, N_control_=11,348), there were no GWS loci. Meta-analysis of the male and female sex-stratified GWAS, including both sexes in the meta-analysis, yielded eight significant loci (Table 2, Supplemental Figure 5), with five significant only in that analysis. Of all the loci significant in the sex-stratified meta-analyses, *POM121L2*\*rs4559087 was the only one with a concordant direction of effect (beta having the same sign) across all four ancestral groups.

### Gene-based tests

Using MAGMA^12^, 15 genes were significantly associated with StimUD in the EUR meta-analysis, reaching a Bonferroni-corrected threshold of *P*=2.63×10^−6^ (Table 3). In the multi-ancestry meta-analysis, six genes were significantly associated with StimUD, reaching a corrected threshold of *P*=2.62×10^−6^ (Table 4). MAGMA found the “behavioral response to ethanol” pathway to be enriched for genes identified in the multi-ancestry meta-analysis (Bonferroni-corrected *P*=0.0054). In the sex-stratified meta-analysis in EUR (males and females together), eleven genes were found to be significantly associated with StimUD, reaching a corrected threshold of *P*=2.63×10^−6^ (Table 5). There were 22 unique genes implicated in the three MAGMA analyses.

**Table 3:** Genes significant in the MAGMA gene-based analysis of the EUR meta-analysis.

| Gene | Chr | Start | Stop | N SNPs | N | Z | P |
| --- | --- | --- | --- | --- | --- | --- | --- |
| *DRD2* | 11 | 113280318 | 113346413 | 258 | 63833 | 6.0804 | 5.99E-10 |
| *SPATS2L* | 2 | 201170604 | 201346986 | 623 | 63280 | 6.0707 | 6.37E-10 |
| *USP19* | 3 | 49145479 | 49158371 | 22 | 57086 | 5.1496 | 1.30E-07 |
| *IREB2* | 15 | 78729773 | 78793798 | 201 | 64077 | 5.029 | 2.47E-07 |
| *TCTA* | 3 | 49449639 | 49453908 | 9 | 69082 | 4.9829 | 3.13E-07 |
| *CCDC36* | 3 | 49235861 | 49295537 | 144 | 60364 | 4.9103 | 4.55E-07 |
| *RP11-3B7.1* | 3 | 49297518 | 49298744 | 5 | 65602 | 4.9048 | 4.68E-07 |
| *ANKK1* | 11 | 113258513 | 113271140 | 61 | 62157 | 4.7861 | 8.50E-07 |
| *LY75-CD302* | 2 | 160628362 | 160761221 | 772 | 62084 | 4.7683 | 9.29E-07 |
| *LY75* | 2 | 160628362 | 160761260 | 772 | 62084 | 4.7683 | 9.29E-07 |
| *C3orf84* | 3 | 49215065 | 49229291 | 37 | 59427 | 4.7121 | 1.23E-06 |
| *TYW5* | 2 | 200794698 | 200820459 | 72 | 63309 | 4.7029 | 1.28E-06 |
| *C2orf69* | 2 | 200775979 | 200820658 | 149 | 61758 | 4.6637 | 1.55E-06 |
| *BSN* | 3 | 49591922 | 49708978 | 260 | 61888 | 4.5689 | 2.45E-06 |
| *C2orf47* | 2 | 200820040 | 200873263 | 147 | 62262 | 4.559 | 2.57E-06 |

**Table 4:** Genes significant in the MAGMAgene-based analysis of the multi-ancestry meta-analysis.

| Gene | Chr | Start | Stop | N SNPs | N | Z | P |
| --- | --- | --- | --- | --- | --- | --- | --- |
| *DRD2* | 11 | 113280318 | 113346413 | 298 | 79991 | 6.0081 | 9.38E-10 |
| *SPATS2L* | 2 | 201170604 | 201346986 | 651 | 86908 | 5.4056 | 3.23E-08 |
| *ANKK1* | 11 | 113258513 | 113271140 | 69 | 79982 | 4.877 | 5.38E-07 |
| *CHRNA5* | 15 | 78857862 | 78887611 | 146 | 74796 | 4.8451 | 6.33E-07 |
| *IREB2* | 15 | 78729773 | 78793798 | 259 | 75083 | 4.7278 | 1.13E-06 |
| *HS6ST3* | 13 | 96743093 | 97485671 | 2857 | 78106 | 4.5607 | 2.55E-06 |

**Table 5:** Genes significant in the MAGMA gene-based analysis of the EUR sex-stratified meta-analysis.

| Gene | Chr | Start | Stop | N SNPs | N | Z | P |
| --- | --- | --- | --- | --- | --- | --- | --- |
| *DRD2* | 11 | 113280318 | 113346413 | 225 | 84191 | 5.1559 | 1.26E-07 |
| *C3orf84* | 3 | 49215065 | 49229291 | 33 | 84191 | 5.0458 | 2.26E-07 |
| *TCTA* | 3 | 49449639 | 49453908 | 9 | 84191 | 4.9245 | 4.23E-07 |
| *CCDC36* | 3 | 49235861 | 49295537 | 128 | 84191 | 4.8876 | 5.10E-07 |
| *FTO* | 16 | 53737875 | 54155853 | 1601 | 84191 | 4.8402 | 6.49E-07 |
| *CCDC71* | 3 | 49199968 | 49203754 | 7 | 84191 | 4.8347 | 6.67E-07 |
| *LAMB2* | 3 | 49158547 | 49170551 | 22 | 84191 | 4.6724 | 1.49E-06 |
| *USP4* | 3 | 49315264 | 49378145 | 119 | 84191 | 4.6377 | 1.76E-06 |
| *RP11-3B7.1* | 3 | 49297518 | 49298744 | 5 | 84191 | 4.6117 | 2.00E-06 |
| *USP19* | 3 | 49145479 | 49158371 | 22 | 84191 | 4.6096 | 2.02E-06 |
| *NICN1* | 3 | 49460379 | 49466759 | 11 | 84191 | 4.5645 | 2.50E-06 |

Using gene set enrichment analysis as implemented in FUMA^13^, the set of genes at 3p21 was enriched for the 22 unique genes significant in the above analyses (Bonferroni-corrected *P*=3.28×10^−10^). Other gene sets significantly enriched for these genes are found in Supplemental Table 1.

### Heritability and genetic correlations

Among EUR datasets, SNP heritability was statistically significant in MVP, AOU, FinnGen, and iPSYCH 1 (Supplemental Table 2), so we evaluated genetic correlations between these datasets. Heritability *Z*-scores ranged from 2.1 (iPSYCH 1) to 9.6 (MVP). Genetic correlations (r_g_) ranged from 0.64 (s.e.=0.12, *P*=2.19×10^−7^) between AOU and FinnGen to 0.80 (s.e.=0.26, *P*=0.0017) between AOU and iPSYCH 1; the r_g_ between FinnGen and iPSYCH 1 was not significant, likely because of limited polygenic signal due to small sample size in iPSYCH 1. We did not observe significant heritability in the remaining smaller EUR datasets and did not evaluate their genetic correlations.

The EUR and AFR meta-analyses had statistically significant SNP heritability (Supplemental Table 3). For EUR, liability scale h^2^=6.1%, s.e.=0.0042, *Z*-score=14.5, population prevalence=0.003.^3,14^ There was no evidence of population stratification or confounding (LDSC intercept=1.02±0.0074, ratio=0.080±0.033). Covariate LDSC (cov-LDSC), using genome-wide principal components (PCs) to estimate LD scores in admixed populations, was used to calculate SNP heritability for AFR and AMR.^15^ For AFR, liability scale h^2^=2.39%, s.e.=0.0067, *Z*-score=3.57, population prevalence=0.001, intercept=1.02±0.007, ratio=0.38±0.12. This relatively high ratio is likely the result of shorter-range linkage disequilibrium and smaller sample size in AFR yielding increased variance in the estimate.^15^ AMR heritability was not significant.

EAS heritability was relatively high (h^2^=40.1%, s.e.=0.15, *Z*-score=2.76); this high point estimate may be an artifact of the small sample size (N_total_=1,015, N_eff_=597). Heritability of StimUD in the individual AFR and AMR datasets that were meta-analyzed is found in Supplemental Table 4.

The genetic correlations for StimUD across ancestries were moderate: between EUR and AFR, r_g_=0.55 (genetic impact, s.e.=0.14, two-tailed *P*=1.46×10^−4^); between EUR and AMR, r_g_=0.63 (genetic impact, s.e.=0.14, *P*=8.45×10^−6^). The genetic correlation between EUR and EAS was not significant.

In the EUR sex-stratified meta-analysis, males had a liability scale h^2^=5.6%, s.e.=0.0051, *Z*=10.9, population prevalence=0.003. Females had a liability scale h^2^=12.6%, s.e.=0.0221, Z=5.7, population prevalence=0.003. Genetic correlation between the male and female meta-analyses was high (r_g_=0.94, *P*=1.21×10^−17^).

We calculated genetic correlations with StimUD for 2,945 traits in EUR (Figure 1, Supplemental Table 5), which revealed widespread overlap with psychiatric and medical traits.^16^ After multiple testing correction (*P*=1.70×10^−5^), 562 genetic correlations were significant. The strongest positive correlations were for opioid use disorder (OUD) (r_g_=1.01, *P*=4.40×10^−107^), viral hepatitis C (r_g_=0.99, *P*=5.80×10^−119^), CocUD (r_g_=0.95, *P*=3.35×10^−121^), and cannabis use disorder (CanUD) (r_g_=0.94, *P*=5.43×10^−237^). Although comorbidity information was not available for all cohorts, 44% of MVP StimUD cases and 41% of AOU StimUD cases also had an OUD diagnosis; 64% of MVP StimUD cases and 35% of AOU StimUD cases also had a CanUD diagnosis. The strongest negative correlations were with smoking status: never (r_g_=-0.61, *P*=3.31×10^−87^), age of first sexual intercourse (r_g_=-0.60, *P*=4.02×10^−99^), and age started oral contraceptive pill (r_g_=-0.52, *P*=5.11×10^−21^). There were high correlations between StimUD and lifetime use of methamphetamine (r_g_=0.82, *P*=8.38×10^−56^) and prescription stimulants (r_g_=0.75, *P*=5.86×10^−33^).

**Figure 1.**
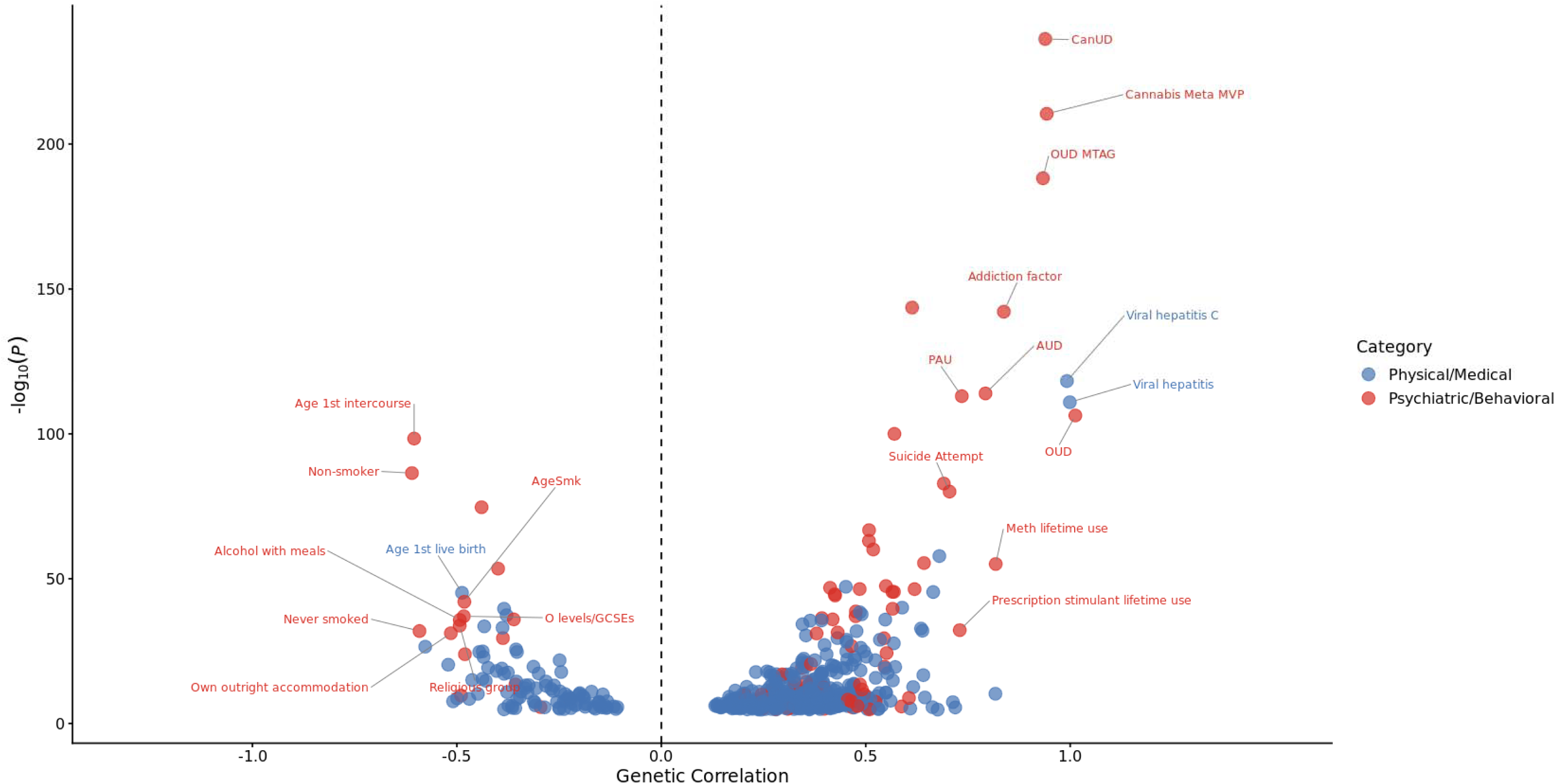
Genetic correlations between StimUD and psychiatric, behavioral, and medical traits. Genetic correlations between StimUD and other traits were estimated using linkage disequilibrium score regression (LDSC). The x-axis shows the estimated genetic correlation (r_g_) with StimUD, and the y-axis shows the strength of evidence for association expressed as -log_10_(*P*). Each point represents a trait. Points for psychiatric/behavioral traits are red and those for physical/medical traits are blue. Traits with the strongest associations are labeled. PAU, problematic alcohol use. AUD, alcohol use disorder. OUD, opioid use disorder.

Using cov-ldsc^15^, genetic correlations were calculated for StimUD in AFR and five other traits in AFR (depression^17^, anxiety^18^, CanUD^19^, CocUD, and OUD^20^), the only summary statistics with significant heritability available in AFR. StimUD had a strong positive correlation with CocUD (r_g_=0.913, *P*=1.17×10^−10^) and CanUD (r_g_=0.830, *P*=3.61×10^−7^). No significant genetic correlations were found in AMR or EAS.

### Pleiotropy analyses

We used MiXeR to quantify the polygenic overlap between StimUD and other psychiatric disorders (Supplemental Tables 6, 7).^21^ MiXeR defines an influential variant as one affecting the trait. Some influential variants are shared between traits and some are specific to a single trait. OUD had 273±498 influential variants specific to OUD and 5,348±807 shared with StimUD. The low point estimate and high standard error suggest that the true number of OUD-specific variants is small. Three models were retained: StimUD-anorexia, StimUD-schizophrenia, and StimUD-bipolar disorder, i.e., MiXeR’s causal mixture model, in which only certain variants influence the phenotype, should be retained only for these three trait pairs. Other traits’ relationships were adequately described with genetic correlations calculated by LDSC using the infinitesimal model, in which every variant influences the phenotype.

### Local genetic correlations

Using LAVA, we identified 78 local genetic correlations between StimUD and other psychiatric traits (Supplemental Table 8), adjusting the *P*-value with an FDR of 5%. Among others, there were local genetic correlations at *CADM2* (3:84698481-85807679) with problematic alcohol use (ρ=0.87, *P_adj_*=0.0066), ADHD (ρ=1, *P_adj_*=0.0099), CanUD (ρ=0.94, *P_adj_*=0.014), and cannabis lifetime use (ρ=0.73, *P_adj_*=0.018).

### Mendelian randomization

After multiple testing correction (28 tests, *P*<0.0018), attention-deficit/hyperactivity disorder (ADHD) genetic liability had a unidirectional causal effect on StimUD (corrected β=0.15, *P*=2.7×10^−6^, *P* of difference between observed and corrected values=4.1×10^−7^). StimUD genetic liability had unidirectional causal effects on neuroticism (observed β=0.68, *P*=0.0014) and stroke (corrected β=0.54, *P*=0.0010, *P_diff_*=0.0016). Anxiety and depression genetic liabilities had bidirectional causal effects: with anxiety as exposure and StimUD as outcome, corrected β=0.44, *P*=1.1×10^−14^, *P_diff_*=3.7×10^−10^; with StimUD as exposure and anxiety as outcome, corrected β=0.99, *P*=2.2×10^−5^, *P_diff_*=0.00011; with depression as exposure and StimUD as outcome, corrected β=0.22, *P*=6.9×10^−21^, *P_diff_*=9.7×10^−19^; with StimUD as exposure and depression as outcome, observed β=0.70, *P*=0.0016 (Supplemental Table 9).

### Transcriptome-wide association study

Using summary statistics from the EUR meta-analysis, we imputed gene expression for autosomes, identifying 44 gene-based associations with TWAS *P*-values <8.7×10^−7^ (0.05/57,337 tests), 19 of which (12 unique genes) survived permutation testing (*P*<0.05) (Supplemental Table 10). For the loci on chromosomes 3 and 9 with multiple TWAS-associated genes, conditional and joint analyses identified jointly significant genes. On chromosome 3, the expression of *GPX1* in the caudate was jointly significant (*P*=1.60×10^−9^), indicating that the same genetic variant(s) affect both *GPX1*’s expression in the caudate and StimUD. After conditioning on the predicted expression of *GPX1*, *QARS1*\*rs4955426, from the EUR meta-analysis, was not significant. On chromosome 9, the expression of *PRPS1P2* in the hippocampus was jointly significant (*P*=3.00×10^−7^).

### Fine-mapping

For loci with multiple TWAS-associated genes, fine-mapping identified those whose expression is most likely to be causally associated (PIP>0.5) with StimUD (Supplemental Table 11).

We also fine-mapped each GWS locus from the EUR meta-analysis. The SNP with the highest PIP overall, rs1543442 (PIP=0.825), in a credible set of 12 SNPs, maps to the 3′UTR of *SNAI1* (chromosome 20). Other SNPs with 0.4<PIP<0.5 are listed in Supplemental Table 12. We extracted functional annotations for the lead SNPs in these credible sets from a dataset of ∼19 million UK Biobank SNPs with 0.005<MAF<0.050 based on the baseline-LF (low-frequency) model (Supplemental Table 13).^22^

### Colocalization analyses

We tested the colocalization of StimUD, brain expression quantitative trait loci (eQTLs)^23^, and brain protein quantitative trait loci (pQTLs)^24^ using HyPrColoc.^25^ Four loci had posterior probability (*P_R_P_A_*)>0.7 that StimUD and brain QTLs colocalize there (Supplemental Table 14).

### Partitioned SNP-based heritability

Partitioned LDSC was used to assign the heritability of StimUD by functional category and cell type.^26^ Of 53 functional categories, Lindblad-Toh conserved noncoding elements^27^, or noncoding regions conserved across mammalian species, showed significant enrichment, with 2.57% of SNPs in the EUR meta-analysis explaining 37.0% of StimUD heritability (*P*=3.63×10^−7^). In the cell-type analysis, neurons were significantly enriched (coefficient=9.97×10^−9^, s.e.=2.76×10^−9^, *P*=0.00015, testing three cell types for enrichment, cutoff *P*<0.05/3).^28^ In the GTEx brain region analysis (0.05/13 tests), cerebellum was significantly enriched (coefficient=9.01×10^−9^, s.e.=3.28×10^−9^, *P*=0.0030).^29^

### Polygenic risk scores

AOU (Table 1) was used as the target dataset for a polygenic risk score (PRS) analysis of StimUD in EUR (N_case_=3,388, N_control_=15,453), AFR (N_case_=2,271, N_control_=9,992), and AMR (N_case_=1,821, N_control_=7,918). Leave-one-out meta-analyses were performed with the remaining summary statistics to produce training datasets. Comparing the fifth quintile of the PRS distribution to the first, PRS were significantly associated with StimUD case status in EUR (odds ratio (OR)=3.28, 95% confidence interval (CI)=2.89-3.71) and AMR (OR=2.01, 95% CI=1.71-2.37), but not in AFR (OR=1.04, 95% CI=0.902-1.20) (Supplemental Table 15).

### Phenome-wide association study

In a phenome-wide association study, we tested the association of specific variants GWS in any of our analyses with other EUR phenotypes, using a threshold of *P*=5.26×10^−8^. We found strong associations between *CHRNA5*\*rs55781567 and smoking traits (lowest *P*=1.03×10^−206^), *ESR1*\*rs3757323 and age at first intercourse (lowest *P*=1.20×10^−37^), *BARHL2*\*rs1526480 and age at first intercourse (lowest *P*=9.30×10^−20^), *FTO*\*rs8057044 and coffee intake (*P*=5.80×10^−21^), and *FTO*\*rs8057044 and hypertension (lowest *P*=1.00×10^−14^) (Supplemental Table 16).

## Discussion

In the current largest GWAS on StimUD, we identified six EUR, one AFR, and five multi-ancestry loci (identified only in the multi-ancestry meta-analysis). Two loci were identified in a male-only GWAS of EUR, with another five identified only in a sex-stratified meta-analysis of EUR, for a total of 19 loci. Three of these loci have, to our knowledge, not previously been associated with any substance use or use disorder (SUD) phenotype, revealing novel biological contributors to this common psychiatric disorder, a leading cause of the U.S. overdose epidemic. Novel loci include *GALNTL4*\*rs11021814 and *CNTN1*\*rs111410321, both in the EUR sex-stratified meta-analysis, and *TRNAQ21*\*rs6915987, in the EUR male-only meta-analysis. We also identified loci in *DRD2*, *CHRNA5*, *FTO*, and *ESR1*, genes that have previously been implicated in studies of SUDs and psychiatric traits, including ADHD.

*DRD2* variants have been previously associated to numerous SUD and psychiatric traits. *DRD2*\*rs5794864 (intronic), significant in EUR (*P*=7.32×10^−10^) and the multi-ancestry meta-analysis (*P*=1.41×10^−8^), was previously identified in a GWAS of alcohol use.^30^ The D2 dopamine receptor, encoded by *DRD2*, is a postsynaptic G-protein-coupled receptor on dopaminergic neurons that is central to the mesocorticolimbic reward pathway and thus relevant to SUDs, especially StimUD and CocUD, and other reward-seeking behavior^31–33^, because a key biological effect of stimulants is increased synaptic availability of dopamine. Multiple positron emission tomography studies have shown that individuals with StimUD have fewer D2 receptors in striatum.^33–37^ Lower D2 dopamine receptor availability is associated with impulsivity^36^, preference for immediate rewards^35^, drug-biased choice^38^, and risk of relapse.^39^ Studies in primates^40,41^ and rats^42^ suggest that reduced D2 receptor availability predisposes to cocaine self-administration. Variants mapping to *DRD2* have been repeatedly identified in GWAS of shared liability to addiction^43–45^, as well as of drinks per week^30,46–48^, alcohol use disorder^49–53^, alcohol consumption^54,55^, cigarettes per day^30,48^, smoking cessation^30,48,56^, smoking status^56^, CanUD^19^, OUD^20,52^, and other psychiatric and neurodevelopmental traits including schizophrenia^57–59^ and ADHD.^60–63^ *DRD2*\*rs4938017 (intronic), another variant significant in EUR and the multi-ancestry meta-analysis, is an eQTL of tetratricopeptide repeat domain 12 (*TTC12*), ankyrin repeat and kinase domain containing 1 (*ANKK1*), and *RP11-159N11.4*.^64^ Expression levels of *RP11-159N11.4* have been associated with neuroticism^65^, loneliness^66^, and other traits related to low mood.

*CHRNA5*\*rs55781567, significant in the multi-ancestry meta-analysis (*P*=3.27×10^−9^), was identified in studies of lung cancer and other pulmonary traits.^67,68^ Other SNPs on *CHRNA5* are associated with nicotine-related traits^48,69,70^ and CocUD.^71,72^ Nicotinic acetylcholine receptor subunit alpha-5, encoded by *CHRNA5,* is expressed on dopaminergic neurons in the ventral tegmental area and nucleus accumbens. Like the D2 dopamine receptor, it plays a key role in the reward pathway.^73^ Other genes in the same 15q25 cluster, including those encoding other subunits of the receptor, have been linked to ADHD.^74–76^ *CHRNA5*\*rs55781567 is also an eQTL for *IREB2*, *PSMA4*, *CTSH*, and *AGPHD1*.^64^ In a study of miRNA as a regulator of methamphetamine-induced behavioral sensitization in mice, IREB2 protein was upregulated in the nucleus accumbens following methamphetamine treatment.^77^ *PSMA4* is at the site of a local genetic correlation between smoking behavior and schizophrenia.^77–79^ Its expression in the cerebellum was significant in our TWAS (permutation *P*=0.00177, *Z*=-5.05). Phase II trials of a selective agonist of the neuronal acetylcholine receptor, pozanicline, have been conducted for the treatment of ADHD; several other agents currently approved for myasthenia gravis also act on this receptor (Supplemental Table 17).^80^

In addition to *DRD2*, 17 other genes identified in our meta-analyses, MAGMA analyses, and TWAS have been GWS in analyses of ADHD (Supplemental Table 18). As shown for the first time in this study, StimUD and ADHD are associated to different SNPs in the same or closely related genes. Taken together, this result, the role of the reward pathway in both disorders, and the use of prescription stimulants as a treatment for ADHD suggest a causal relationship between the two disorders. Previous studies have shown that prescription stimulant treatment of ADHD may be protective against SUDs^81–83^, but more recent studies have found no association once confounders are taken into account.^84^ Our bioinformatic analyses suggest a partly causal association. MR analysis revealed that genetic liability to ADHD had a significant effect on StimUD. If genetic liability to ADHD predisposes to StimUD, treating ADHD may mitigate the risk of developing StimUD or another SUD, consistent with findings from earlier studies.^81–83^ For example, if ADHD were adequately treated with prescription stimulants, this might make self-medication with methamphetamine or diverted prescriptions less likely. There was a high r_g_ between StimUD and other SUDs. Using a constraint test implemented in GenomicSEM^85^, the StimUD-ADHD r_g_ was lower than those for StimUD and SUDs, but significantly higher than between StimUD and other psychiatric disorders (Supplemental Table 5). Thus, there is some evidence for causation.

There are no FDA-approved drug treatments for StimUD^86^, but there are many approved drugs that act on the D2 dopamine and nicotinic acetylcholine receptors, as well as a current randomized clinical trial of mirtazapine for methamphetamine use disorder.^87^ Additional druggable targets include proteins encoded by *FTO* and *ESR1*, in addition to other less-well-known protein targets (Supplemental Tables 17, 19). Of particular interest are the genes identified in brain pQTL colocalization analyses, *CWF19L1*, *TMX2-CTNND1*, and *TANC2* (Supplemental Table 14), because if a protein’s expression level is associated with disease, then pharmacologically targeting (e.g., inhibiting) that expression could be therapeutic. At least seven currently-approved drugs act on *FTO* (Supplemental Table 17). *FTO*\*rs8057044, significant in the EUR male-only meta-analysis and the EUR sex-stratified meta-analysis of males and females, was previously identified in GWAS of body mass index and metabolic syndrome.^88–90^ Its direction of effect was the same in males and females, but the magnitude was greater in males (β=-0.080 in males, β=-0.041 in females), suggesting that this variant has a greater effect on the phenotype in males. Another SNP at this locus, *FTO*\*rs56137030, previously identified in studies of substance-related disorders^44,45^ and nicotine dependence^91^, was GWS in males but not the sex-stratified meta-analysis, and similarly had β=-0.082 in males, but only β=-0.013 in females. Other SNPs in *FTO* have been associated with the addiction risk factor^44^, cigarettes per day^88^, alcohol use and use disorder^45,51–55,92,93^, opioid dependence^20,94,95^, coffee or tea consumption^50,96,97^, bipolar disorder^98^, and schizophrenia.^99,100^ Thus, *FTO* is ripe for drug development or repurposing.

*ESR1,* encoding the estrogen receptor 1, is highly expressed in the hippocampus, where it modulates synaptic plasticity related to memory^101,102^, and the hypothalamus, where it regulates reproduction and homeostasis.^103^ *ESR1*\*rs3757323, significant in the multi-ancestry meta-analysis (*P*=3.30×10^−8^), has been associated with suicidal ideation^104^ and major depressive disorder^105^, and is an eQTL of coiled-coil domain containing 170 (*CCDC170*), which has been identified in GWAS of executive function in ADHD^106^, number of sexual partners^46,107^, suicide attempt^108^, depression^65^, and risk-taking behavior.^109^ Other SNPs in *ESR1* were identified in GWAS of methamphetamine-induced psychosis^110^, executive function in ADHD^106^, SUDs^43^, mood disorders^45^, alcohol dependence^111^, anxiety^18,112^, suicide^113^, educational attainment^50,114–117^, depression^105,118,119^, and externalizing behavior.^120^

This study has limitations. First, phenotypic heterogeneity among cohorts is a consideration. MVP, All of Us, FinnGen, iPSYCH 1, iPSYCH 2, and MGBB use ICD codes from an EHR, Yale-Penn and COGA use diagnoses made using semi-structured interviews, and QIMR participants completed structured questionnaires. There is also heterogeneity in the selection of controls.

Second, due to the nature of ICD coding, we lack specific information on the type of stimulant taken, which could include illicit methamphetamine, diverted prescription stimulants, or caffeine, although it is unlikely that caffeine users often receive StimUD diagnoses. Finally, sample sizes in AFR, AMR, and EAS were much smaller than in EUR, which limited both GWAS discovery and, for AFR, post-GWAS analysis.

This is the largest genome-wide analysis of StimUD to date, including more than 700,000 participants from four ancestral groups and several international cohorts. We uncovered novel genetic associations, identified associations to *DRD2*, *CHRNA5*, *ESR1*, and *FTO* of particular and immediate biological interest, and used the results to investigate genetic relationships between StimUD and other psychiatric traits. MR revealed bidirectional causal relationships between StimUD, anxiety, and depression, and unidirectional causal effects of ADHD on StimUD and StimUD on neuroticism. The unidirectional estimated causal effect of StimUD on stroke provides one example where StimUD may have clinical consequences beyond its psychiatric comorbidities. The ancestral diversity of MVP, AOU, and several smaller cohorts allowed us to investigate StimUD in non-EUR samples. Genetic correlation analysis of StimUD in AFR showed a high genetic correlation with CanUD and CocUD in AFR, as we found in EUR, while PRS analysis showed that polygenic risk for StimUD predicted StimUD in AMR, just as in EUR. The large size of MVP and AOU allowed for sex-stratified analyses in EUR in these cohorts, revealing additional GWS loci that may have differential effects by sex. Gene-based, transcriptomic, and colocalization analyses prioritized additional genes (Tables 3-5; Supplemental Tables 8, 10, 11, 12, 14), some of which have potential for drug development or repurposing (Supplemental Tables 17, 19, 20) or have been identified in GWAS of ADHD (Supplemental Table 18). The overlap between StimUD and other disorders and the many drug targets identified offer multiple areas for future research.

## Methods

### Inclusion and ethics statement

Researchers from the Collaborative Study on the Genetics of Alcoholism (COGA)^121–123^, QIMR Berghofer (QIMR)^124,125^, Lundbeck Foundation Initiative for Integrative Psychiatric Research (iPSYCH)^29,126,127^, Mass General Brigham Biobank (MGBB)^128^, the University of New South Wales Sydney^129,130^, Washington University in St. Louis^131^, and the Yale-Penn Study^132–134^ contributed summary statistics to these meta-analyses. All studies were approved by local institutional research boards and ethics review committees. The MVP was approved by the Veterans Affairs Central Institutional Review Board. Additional information can be found in the Supplementary Material.

### Cohorts

New GWAS were conducted in all datasets except FinnGen, where summary statistics were downloaded from https://console.cloud.google.com/storage/browser/finngen-public-data-r12/summary_stats. MVP, FinnGen, AOU, and QIMR analyses included the X chromosome. The Yale-Penn cohort comprises three datasets, Yale-Penn 1, Yale-Penn 2, and Yale-Penn 3, in which GWAS were run separately; the iPSYCH cohort comprises two datasets, iPSYCH 1 and iPSYCH 2, in which GWAS were run separately, for a total of 13 datasets in the EUR meta-analysis (Table 1). Details of phenotyping, genotyping, imputation, and GWAS for each cohort are found in the Supplementary Material. Table 1 provides a case-control and sex breakdown for each dataset, Supplemental Table 21 summarizes phenotype definitions across cohorts, and Supplemental Tables 22 (ICD-9-CM) and 23 (ICD-10-CM) define the ICD codes used.

### LDSC and SNP-based heritability

To calculate liability-scale SNP heritability, we used LDSC^16^ for EUR and EAS, cov-LDSC^15^ for AFR and AMR, and Popcorn for between-ancestry comparisons^135^, using a lifetime population prevalence of StimUD of 0.3% for EUR^3,14^, 0.1% for AFR, 0.1% for AMR, and 0.1% for EAS.^3^ 3.49% of individuals met criteria for StimUD in the EUR meta-analysis, 18.1% in the AFR meta-analysis, 18.9% in the AMR meta-analysis, and 17.9% in the MVP EAS sample. The proportion of affecteds was much higher for AFR, AMR, and EAS than for EUR because we matched cases and controls by age and sex in a ratio of approximately 1:4 (see Supplementary Material). The higher proportion of controls to cases in EUR was largely because of FinnGen, from which only summary statistics with a large predominance of controls were available. LDSC requires a reference panel to account for LD between some variants. LDSC uses the 1000 Genomes (1KG) EUR and EAS reference panels, but identifying a reference for admixed ancestries is challenging. Cov-LDSC uses a 1KG reference panel^136^ to calculate within-ancestry PCs to use as covariates to adjust its estimate of SNP heritability in that ancestry. Popcorn uses 1KG reference panels to calculate cross-ancestry covariance scores for EUR and AFR, EUR and AMR, and EUR and EAS. LDSC was used both locally and via CTG-VL^137^ to estimate genetic correlation between StimUD and 2,945 traits (2,871 from CTG-VL and 74 in-house) in EUR. Cov-LDSC was used with AFR and AMR to estimate genetic correlation between StimUD and other traits for which summary statistics in that ancestry were available. As with EUR, LDSC was used to calculate heritability and genetic correlations for EAS.

### MAGMA

MAGMA, here implemented in FUMA^13^, tests the joint association of all SNPs in a gene with StimUD.^12^ 1KG reference panels (one for EUR, one for all ancestries) were used.^136^ SNPs from the EUR meta-analysis mapped to 19,040 protein-coding genes and SNPs from the multi-ancestry meta-analysis mapped to 19,002 protein-coding genes.

### Pleiotropy analyses

Disorders were chosen for analysis based on the availability of high-quality summary statistics, either from the Psychiatric Genomics Consortium^138^ or from previously published studies from our lab (the Gelernter lab, Yale School of Medicine).^19,20,53,139^ MiXeR used the Akaike information criterion (AIC) to compare the best bivariate model to that with the least possible polygenic overlap >0, assuming that traits with non-zero genetic correlation have non-zero polygenic overlap and retaining the model with a lower AIC. MiXeR^21^ and LAVA^140^ revealed pleiotropy between StimUD and other psychiatric traits (ADHD, depression, bipolar disorder, anxiety, schizophrenia, anorexia, post-traumatic stress disorder, suicide, obsessive-compulsive disorder, Tourette’s syndrome, cannabis lifetime use, CanUD, problematic alcohol use, and OUD) in EUR. MiXeR yielded the number of causal variants shared with these disorders after excluding the MHC region (6:26000000–34000000). LAVA estimated local genetic correlations with these disorders.

### Mendelian Randomization

Using MRlap^141^, psychiatric traits with SNP-r_g_≥0.4 with StimUD, five-factor personality traits^142^, and somatic traits related to hypertension, given its correlation with prescription stimulant use^143^, were analyzed for bidirectional causal relationships with StimUD in EUR. Only independent GWS variants (*P*<5×10^−8^, LD r^2^<0.05, >500 kB apart) were used as genetic instruments.

### Transcriptome-wide association study

FUSION^144^ was used for a transcriptome-wide association study (TWAS), integrating GWAS results and gene expression data to characterize gene expression patterns associated with StimUD in EUR. TWAS models were trained using GTEx v8 expression data^23^ from 13 regions in adult brain and spinal cord, including amygdala (119 samples, 2,660 genes), anterior cingulate cortex BA24 (135 samples, 3,512 genes), caudate (172 samples, 5,128 genes), cerebellar hemispheres (157 samples, 6,200 genes), cerebellum (188 samples, 7,387 genes), cortex (183 samples, 5,695 genes), frontal cortex BA9 (157 samples, 4,603 genes), hippocampus (150 samples, 3,613 genes), hypothalamus (156 samples, 3,611 genes), nucleus accumbens (181 samples, 5,086 genes), putamen (153 samples, 4,362 genes), spinal cord (115 samples, 3,178 genes), and substantia nigra (100 samples, 2,302 genes). GTEx samples were not ascertained for StimUD. Genes in the models had significant non-zero *cis*-heritability at *P*<0.01. Models underwent five-fold cross-validation by random sampling of 1,000 significantly heritable genes. The *R*^2^ between predicted and true gene expression was >0.01 in each case (*P*<0.01).^19,144^ Wald-type *Z*-scores were generated for each gene in each tissue using a weighted burden test; transcriptome-wide significance meant *P*<8.7×10^−7^ (0.05/57,337 tests).

The 1KG EUR reference panel was used.^136^ Permutation tests were conducted; SNP-gene weights in the models were shuffled 10,000 times to generate null distributions to which the TWAS *Z*-scores were compared. Loci with more than one significant gene were fine-mapped using FOCUS^145,146^, generating a 90% credible set of genes whose transcription levels were causal for StimUD. FOCUS used a database of eQTLs from 13 brain regions.^23^

### Fine-mapping with functional annotations

PolyFun (POLYgenic FUNctionally-informed fine-mapping) and SuSiE (Sum of Single Effects) were used to perform SNP-level fine-mapping.^147,148^ PolyFun uses summary statistics to calculate per-SNP heritability, using it to estimate prior causal probability. We extracted annotations for variants with posterior inclusion probability (PIP) at least 0.5 using 187 functional annotations for approximately 19 million UK Biobank SNPs with 0.005<MAF<0.050.^22^

### Colocalization

We used HyPrColoc^25^ to assess colocalization in EUR of StimUD, eQTLs from basal ganglia, cerebellum, cortex, hippocampus, and spinal cord^149^, and brain pQTLs.^24^ In AFR, we analyzed StimUD and cortex eQTLs; that was the only region available.^149^ The genome was divided into 1,703 approximately independent LD blocks; each was analyzed to yield posterior probabilities (*P_R_P_A_*) of putatively causal SNPs in that region. A *P_R_P_A_* >0.7 was the threshold for colocalization.

### Stratified LDSC

Stratified LD score regression^26^ partitions heritability of StimUD by functional annotation while accounting for LD among markers. The 53 baseline annotations included coding, UTR, promoter, intron, histone marks, open chromatin indicated by DNAse I hypersensitivity site (DHS) regions, chromHMM/Segway predictions, which partition the genome into seven chromatin states, regions conserved in mammals, super-enhancers (large clusters of highly active enhancers), and FANTOM5 enhancers.^26^ There were additionally 205 cell-type-specific annotations derived from histone marks H3K4me1, H3K4me2, H3K9ac, and H3K27ac, 13 brain-region-specific annotations, and three brain-cell annotations.

### Polygenic risk scoring

Leave-one-out meta-analyses were performed excluding AOU from EUR and AFR meta-analyses (excluding AOU from the AMR meta-analysis left only MVP). We conducted PRS analyses using the leave-one-out analyses as the training datasets and AOU EUR, AFR, and AMR as the target datasets. X-Wing^150^ generated SNP weights for EUR, AFR, and AMR. PLINK calculated individual-level PRS in each ancestry. For example, AFR individuals have an EUR-based PRS and an AFR-based PRS. X-Wing additionally calculated weights for each ancestry’s PRS, so that the final PRS for an individual was the weighted linear combination of their PRS for each ancestry.

### Phenome-wide association study

Using specific variants found to be significant in our meta-analyses, we conducted a phenome-wide association study over 50,055 UKB, FinnGen, and European Bioinformatics Institute GWAS datasets hosted by the IEU OpenGWAS project^151,152^ to determine whether these variants were significantly associated with other medical or psychiatric traits.

### Reporting summary

Further information on research design is available in the Nature Portfolio Reporting Summary linked to this article.

## Acknowledgements

This research is based in part on data from the Million Veteran Program, Office of Research and Development, Veterans Health Administration, and was supported by MVP000 as well as award I01BX006482-05. More details regarding this consortium are available in the Supplementary Information. Further support was provided by NIH (grants 3R01DA058862 to J.G., R01DA054869 to J.G., R01DA037974 to J.G., K01DA058807 to J.D., R01AA031176 to D.L., 1K01MH141330 to J.T., U10AA008401 to L.W., R01HG012354 and R01MH130899 to T.G., R01DA054869-05 to H.E., K01DA051759 to E.C.J., K01AA030083 to A.H., R01DA054869 and U10AA008401 to A.A., R01MH137218 to J.W.S., and K08MH135343 to T.M.). D.F.L. is supported by a Career Development Award CDA-2 from the Veterans Affairs Office of Research and Development (1IK2BX005058). H.K. is supported by V.A. grant I01BX004820 and the VISN 4 Mental Illness Research, Education and Clinical Center. This publication does not represent the views of the Department of Veterans Affairs or the United States Government. The Australian Genetics of Depression Study (AGDS) was primarily funded by the National Health and Medical Research Council (NHMRC) of Australia Grant No. 1086683 to N.G.M. N.G.M. was supported by an NHMRC Investigator Grant (No. APP1172990). We are indebted to all of the participants for giving their time to contribute to this study. We wish to thank all the people who helped in the conception, implementation, media campaign, and data cleaning. We thank R. Parker, S. Cross and L. Sullivan for their valuable work coordinating all the administrative and operational aspects of the AGDS project. We would also like to thank the research participants for making this work possible. The Australian Genetics of Bipolar Disorder Study (GBP) data collection was funded and data analysis was supported by the Australian NHMRC (no. APP1138514) to S.E.M. S.E.M. is supported by a NHMRC Investigator Grant (no. APP2025674). We thank the participants for giving their time and support for this project. We acknowledge and thank M. Steffens for her generous donations and fundraising support.

Funding support for the Comorbidity and Trauma Study (dbGAP accession number, phs000277.v1.p1) was provided by NIDA (R01DA17305). Funding support for the Opioid Misuse Study (dbGAP accession number, phs003684.v1.p1) was provided by NIDA (R01DA042620).

D.D. was supported by the Novo Nordisk Foundation (NNF20OC0065561, NNF21SA0072102), the Lundbeck Foundation (R344-2020-1060), the European Union’s Horizon 2020 research and innovation program under grant agreement No. 965381 (TIMESPAN).

The iPSYCH team was supported by grants from the Lundbeck Foundation (R102-A9118, R155-2014-1724, and R248-2017-2003), NIH/NIMH (1R01MH124851-01 to A.D.B.), EU’s Horizon Europe program under grant agreement no. 101057385 (R2D2-MH; to A.D.B.) and the Universities and University Hospitals of Aarhus and Copenhagen. High-performance computer capacity for handling and statistical analysis of iPSYCH data on the GenomeDK HPC facility was provided by the Center for Genomics and Personalized Medicine and the Centre for Integrative Sequencing, iSEQ, Aarhus University, Denmark (grant to A.D.B.).

L.D. is supported by an Australian National Health and Medical Research Council (NHMRC) Investigator Award Level 3 (#2016825). The Australian National Drug and Alcohol Research Centre is supported by funding from the Australian Government Department of Health under the Drug and Alcohol Program.

## Author contributions

S.B., J.D.D., and J.G. designed the study. S.B. and J.G. drafted the manuscript. D.F.L. provided ongoing feedback and refinement of the analytical plan. S.B., T.G., P.W.J., D.L., T.T.M., T.T.N., J.D.T., L.W., and P.L. conducted GWAS on included cohorts. S.B., J.D.D., D.F.L., and J.G. discussed, created, and refined the phenotype in MVP. S.B., J.D.D., and D.F.L. discussed and refined MVP analytic plans. S.B. conducted original analyses. J.D.D., D.F.L., and J.G. supervised original analyses. J.G. supervised the work. All authors critically evaluated and revised the manuscript.

## Data availability

All MVP and meta-analysis summary statistics are made available through dbGAP, https://dbgap.ncbi.nlm.nih.gov, under accession number phs001672.

Meta-analysis summary statistics are available through the Gelernter lab website: https://medicine.yale.edu/lab/gelernter/.

## Code availability

Code for all software packages used in this analysis is publicly available via the citations for each method.

## Competing interests

D.D. has received speaker fees from Takeda and Medice Nordic. H.K. is a member of advisory boards for Altimmune and Clearmind Medicine; a consultant to Sobrera Pharmaceuticals, Altimmune, Lilly, and Ribocure; and the recipient of research funding and medication supplies for an investigator-initiated study from Alkermes and company-initiated studies by Altimmune and Lilly. M.B.S. has in the past 3 years received consulting income from atai Life Sciences, Ananda Scientific, BigHealth, Biogen, Bionomics/Neuphoria, Boehringer Ingelheim, EmpowerPharm, Engrail Therapeutics, Jazz Pharmaceuticals, Karuna Therapeutics, Lundbeck, Lykos Therapeutics, Newleos Therapeutics, Orion Pharma, Otsuka US, PureTech Health, Roche/Genentech, Sage Therapeutics, Seaport Therapeutics, Sensorium Therapeutics, and Transcend Therapeutics. Dr. Stein has stock options in EpiVario, Newleos Therapeutics, and Oxeia Biopharmaceuticals. He has been paid for his editorial work on *Depression and Anxiety* (Editor-in-Chief), *Biological Psychiatry* (Deputy Editor), and *UpToDate* (Co-Editor-in-Chief for Psychiatry). He is on the scientific advisory board of the Brain and Behavior Research Foundation and the Anxiety and Depression Association of America. J.W.S. is a member of the Scientific Advisory Board of Sensorium Therapeutics (with options), has received consulting fees from Tempus, Inc., and has received grant support from Biogen, Inc. J.G. is paid for editorial work for the journal Complex Psychiatry.

## Footnotes

Publisher’s note: Springer Nature remains neutral with regard to jurisdictional claims in published maps and institutional affiliations.

A list of authors and their affiliations appears at the end of the paper.

## Contributor information

Sarah E. Beck

Joel Gelernter

## Supplementary information

The online version contains supplementary material available at […].

## Associated data

*This section collects any data citations, data availability statements, or supplementary materials included in this article*.

### Data availability statement

Data for TWAS models used are available from the Adult Genotype-Tissue Expression Project (GTEx v8), https://gtexportal.org/home/downloads/adult-gtex.

Code for software and packages used in this analysis are all publicly available through the citations for each method as described.

## Notes

### Author Declarations

The Veterans Affairs Central Institutional Review Board gave ethical approval for the Million Veteran Program. The All of Us Institutional Review Board gave ethical approval for All of Us. The iPSYCH study was approved by the Scientific Ethics Committee (SEC) in the Central Denmark Region (Case No 1-10-72-287-12) and the Danish Data Protection Agency.

