## Supplementary Material for "Multi-ancestry genome-wide association study and meta-analysis of stimulant use disorder reveals biology and relationships to other psychiatric disorders"

**Supplementary Information**

- **Supplementary Methods**

1. Inclusion and ethics
2. Phenotyping
3. Genotyping

- **Supplementary Figures**
  1. Manhattan and Q-Q plots of StimUD GWAS meta-analysis results in EUR
  2. Manhattan and Q-Q plots of StimUD GWAS meta-analysis results in AFR
  3. Manhattan and Q-Q plots of StimUD GWAS multi-ancestry meta-analysis results
  4. Manhattan and Q-Q plots of StimUD GWAS male-only meta-analysis results in EUR
  5. Manhattan and Q-Q plots of StimUD GWAS sex-stratified meta-analysis results in EUR
- **VA Million Veteran Program Core Acknowledgement**
- **References**

**Please refer also to Table 1 in the main article for study demographics.**

**Inclusion and ethics**

The iPSYCH study was approved by the Scientific Ethics Committee (SEC) in the Central Denmark Region (Case No 1-10-72-287-12) and the Danish Data Protection Agency. In accordance with Danish legislation, the SEC waived the need for specific informed consent in biomedical research based on existing biobanks. iPSYCH was initially approved by the SEC in 2012, with subsequent amendments in 2013, 2015, and 2018. More details can be found at https://ipsych.dk/en/data-security/health-research-and-ethical-approval. New Danish legislation (effective from January 2024) introduces the possibility for participants to opt out of studies that are exempt from active informed consent. After consulting with the SEC and patient organizations, iPSYCH contacted all ~140,000 participants in the iPSYCH cohort in June 2025 and offered the possibility to opt out of new genetic studies initiated henceforth. Overall, 1.8% of the iPSYCH participants chose to opt out, and their data were deleted from the active research database. Data included in finalized and ongoing studies were not removed.

**Phenotyping in MVP, AOU, and Yale-Penn**

Using the Veterans Affairs (VA) Informatics and Computing Infrastructure, we searched de-identified electronic health records (EHRs) in version 23.1 (April 29^th^, 2024) of the MVP (N=1,016,584) to find individuals with International Classification of Diseases (ICD) codes for amphetamine and/or stimulant abuse and/or dependence (Supplemental Table 21, Supplemental Table 22, Supplemental Table 23). ICD-9-CM (using “amphetamine”) and ICD-10-CM (using “stimulant”) codes were used. Visits occurred between September 4^th^, 1992 and September 30^th^, 2023. Cases had at least one inpatient or outpatient visit to a VA facility with a diagnosis of StimUD. Controls had no ICD codes for StimUD. For all cohorts including MVP, a minimum of 50 cases was needed for analysis. Similarly, in the AOU Researcher Workbench (N=414,830), we searched de-identified EHRs in version 8 (February 3^rd^, 2025) for those with at least one inpatient or outpatient visit with an ICD-9-CM or ICD-10-CM code for amphetamine and/or stimulant abuse and/or dependence (Supplemental Tables 17 and 18). Visits occurred between January 8^th^, 1981 and October 1^st^, 2023. Controls were all those with EHR data who did not meet case criteria. To reduce noise, controls were matched with cases in a ratio of approximately 4:1. In both MVP and AOU, to remove relatives while retaining as many cases as possible, an algorithm was used to preferentially retain the case when a case and a control were related. Pairs of individuals with a kinship coefficient >0.0884 were defined as related. The Yale-Penn cohort (comprising Yale-Penn 1, 2, and 3; N=11,345 with genetic data) has been described previously^1-3^; cases met the DSM-IV criteria for stimulant abuse or stimulant dependence as derived from the Semi-structured Assessment for Drug Dependence and Alcoholism (SSADDA)^4^, while controls did not. In meta-analyses, we included summary statistics from FinnGen release 12, COGA, QIMR, iPSYCH 1, MGBB, CATS, iPSYCH 2, and OMS.

**Genotyping, imputation, quality control, GWAS, and meta-analysis**

Genotyping and imputation of MVP subjects has been described previously.^5,6^ Genotyping was performed using a customized Affymetrix Axiom Array. Genotype data for biallelic SNPs was imputed using Minimac4^7^ and the African Genome Resources reference panel from the Sanger Institute.^8^ Using the 1000 Genomes (1KG) phase 3 reference panels^9^, complex variants and indels were independently imputed with an approach like that of the UK Biobank.^10^

PLINK 2.0^11^ was used for GWAS in MVP using logistic regression. Covariates were sex, age, and the first ten within-ancestry PCs. Variants with call missingness >20% in the best-guess genotype or Hardy-Weinberg equilibrium (HWE) *P<*5×10^-5^ were excluded, as were alleles with minor allele frequency (MAF) <0.001% and per-individual genotyping rate <90%. MVP was the largest, most diverse cohort (Table 1).

Recruitment, assessment, and whole-genome sequencing in AOU have been previously described.^12^ The details of their genomic quality control are at https://support.researchallofus.org/hc/en-us/articles/29390274413716-All-of-Us-Genomic-Quality-Report, including checks for sex concordance, cross-individual contamination, and coverage (mean 30x). Case counts are found in Table 1. There were 26 EAS cases and 22 SAS cases, below our predefined minimum of 50 cases for inclusion. Controls and cases were defined using ICD codes as above for MVP and matched in a ratio of approximately 4:1 as above. Related individuals were excluded using the same algorithm. Quality control was conducted as in MVP. GWAS was performed in PLINK 2.0 using logistic regression. Covariates were age, sex, and the first six within-ancestry PCs for EUR, the first 14 for AFR, and the first ten for AMR. The number of PCs used was determined by reviewing scree plots (percentage of variance explained by each PC) and choosing the point at which the slope of the plot changed as the cutoff for number of PCs.

From FinnGen, we used summary statistics from a GWAS of StimUD in EUR conducted using data from release 12 (November 4^th^, 2024) for phenotype “Mental and behavioural disorders due to use of other stimulants, including caffeine” (F5_STIMUL) (Supplemental Table 21). Case counts are found in Table 1. Information about release 12 is available at https://finngen.gitbook.io/documentation; details of the phenotype are at https://risteys.finngen.fi/endpoints/F5_STIMUL and in Supplemental Table 21. As previously described, REGENIE^13^ was used to conduct the FinnGen GWAS, using age, sex, first ten within-ancestry PCs, and genotyping batch as covariates.^14,15^

COGA’s genotyping, quality control, ancestry assignment, and imputation have been previously reported.^16^ Briefly, samples were genotyped in five batches using four different arrays: Illumina Human 1M, the Illumina Human OmniExpress 12V1, Illumina Human Omni 2.5M, and the Smokescreen Genotyping Array. SNPs with genotyping rate <95%, minor allele frequency (MAF) <3%, or Hardy-Weinberg equilibrium (HWE) *P*-value <0.0001 were excluded. Genotype data were phased using SHAPEIT2^17^ then imputed using Minimac3^18^ with 1000 Genomes Project data as the reference panel.^9^ Case counts are found in Table 1; phenotype information is in Supplemental Table 21. GWAS was conducted using the R package GWAF^19^ with age, sex, array indicators, and first ten PCs as covariates.

For QIMR, samples (Australian Genetics of Depression Study (AGDS) and Australian Genetics of Bipolar Disorder Study (GBP)) were genotyped using the Illumina Global Screening Array v1 or v2; per-batch imputation QC removed variants with GenTrain score<0.6, MA <0.01, SNP call rate<95% and HWE deviation (*P*<1×10^−6^) before imputation was conducted in TOPMed using the TOPMed-r2 reference panel.^20^ DSM-5 case counts are found in Table 1; phenotype information is in Supplemental Table 21. DSM-5 case status was assessed using the Composite International Diagnostic Interview (CIDI)^21^, based on responses to questions about use of “amphetamine-type stimulants (e.g., ice, speed).” GWAS was conducted in SAIGE v0.44^22^ using sex, age, first ten within-ancestry PCs, and a cohort variable as covariates, and variants were restricted to those with MAF≥0.0001, minor allele count at least 5, and r^2^≥0.1.

Quality control and imputation in iPSYCH1 and iPSYCH 2 have been previously described.^23-25^ IPSYCH 1 was genotyped on the Illumina Infinium PsychChip v1.0; iPSYCH 2 was genotyped on the Illumina Infinium Global Screening Array. Imputation was conducted using Minimac3^7^ and the HRC Release 1.1 reference panel.^26^ Cases were those age 25 or older with an ICD-10 F15.1-9 diagnosis of stimulant use disorder, excluding those with only an acute intoxication diagnosis (F15.0). Controls included those with other psychiatric disorders, in an effort to address bias introduced by other psychiatric disorders by ensuring that cases and controls have similar comorbidities. Case counts are found in Table 1. GWAS was conducted using additive logistic regression in PLINK v1.9 implemented in RICOPILI^27^, and age, sex, and the first ten principal components were included as covariates; only individuals with a subject call rate >0.95 were included, and variants with call rate >0.98, imputation INFO score >0.8, MAF>0.01 and Hardy-Weinberg equilibrium (HWE) *P*>10^−6^ in controls or *P*>10^−10^ in cases were retained for further analysis.

MGBB subjects were genotyped on two Illumina arrays: GSA and MEGA. Imputation was performed on the Michigan Imputation Server^18^ using the 1KG reference panel.^9^ Quality control for the GSA array is described at https://github.com/getian107/MGBB-QC and included variants with INFO r^2^>0.6, MAF>0.005, HWE *P*>1×10^-10^, and SNP-level call rate >0.98. Quality control for the MEGA array is described at https://github.com/Annefeng/PBK-QC-pipeline and included variants with INFO r^2^>0.8, MAF>0.01, HWE *P*>1×10^-10^, and SNP-level call rate >0.98. All samples were restricted to those that had a genotyped variant call rate >0.98. Case counts are found in Table 1 and phenotype information is in Supplemental Table 21. PLINK 2.0^11^ was used for GWAS on variants with MAF>0.05. Covariates included sex, age, and the first 20 within-sample genetic PCs.

In CATS, samples were genotyped using the Illumina Human660W-Quad BeadChip. Pre-imputation QC removed SNPs with HWE *P*<1×10^−6^, MAF<0.01, SNP call rate<95%, or GenomeStudio genotype quality score<0.7. Imputation was performed on the Michigan Imputation Server using the 1000 Genomes Phase 3 reference panel. Case counts are found in Table 1. Cases had DSM-IV stimulant abuse or dependence as assessed by a modified SSAGA^28^; controls were unassessed and population-based. GWAS was conducted using PLINK 2.0^11^ with age, sex, and the first ten within-ancestry principal components as covariates.

In OMS, samples were genotyped using the Illumina Infinium Global Screening Array version 3; pre-imputation QC removed variants with MAF<0.01, SNP call rate<95%, and HWE *P*<1×10^−6^. Imputation was conducted with TOPMed using the TOPMed-r3 reference panel.^20^ Post-imputation, variants were retained with MAF≥0.005 and r^2^≥0.3. Case counts are found in Table 1. Cases had DSM-IV stimulant abuse or dependence as assessed by a modified SSAGA^28^; controls were either opioid-exposed or had OUD. GWAS was conducted using PLINK 2.0^11^ with age, sex, and the first ten within-ancestry principal components as covariates.

Genotyping, imputation, and quality control in Yale-Penn have been previously described.^1-3^ Genotyping was performed using the Illumina HumanOmni1-Quad v1.0 (YP1), the Illumina HumanCore Exome Array (YP2), or the Illumina Multi-Ethnic Global Array (YP3). Imputation was performed with Minimac3^7^ via the Michigan Imputation Server^18^, using HRC^26^ (YP1, YP3) or 1KG^9^ (YP2). GEMMA was used for GWAS to account for relatedness.^29^ PLINK 1.9^11^ was used for quality control with parameters as in MVP above, then GEMMA^29^ was used to calculate a relatedness matrix for GWAS.

EUR cohorts (MVP, AOU, FinnGen, COGA, QIMR, iPSYCH 1, iPSYCH2, MGBB, CATS, OMS, Yale-Penn 1, Yale-Penn 2, Yale-Penn 3) were combined in an effective sample size-weighted GWAS meta-analysis using METAL (Table 1).^30^ N_eff_=4/(1/N_cases_+1/N_controls_). For AFR, OMS, MVP, AOU, and Yale-Penn 1 were meta-analyzed (Yale-Penn 2 and 3 did not have sufficient cases for GWAS). For AMR, MVP and AOU were meta-analyzed. EAS was present in sufficient numbers for analysis only in the MVP. A multi-ancestry meta-analysis including all cohorts was performed. The EUR meta-analysis included 22,564 cases and 624,672 controls, AFR included 7,574 cases and 34,189 controls, AMR included 3,657 cases and 15,698 controls, and the multi-ancestry meta-analysis included 33,977 cases and 675,392 controls.

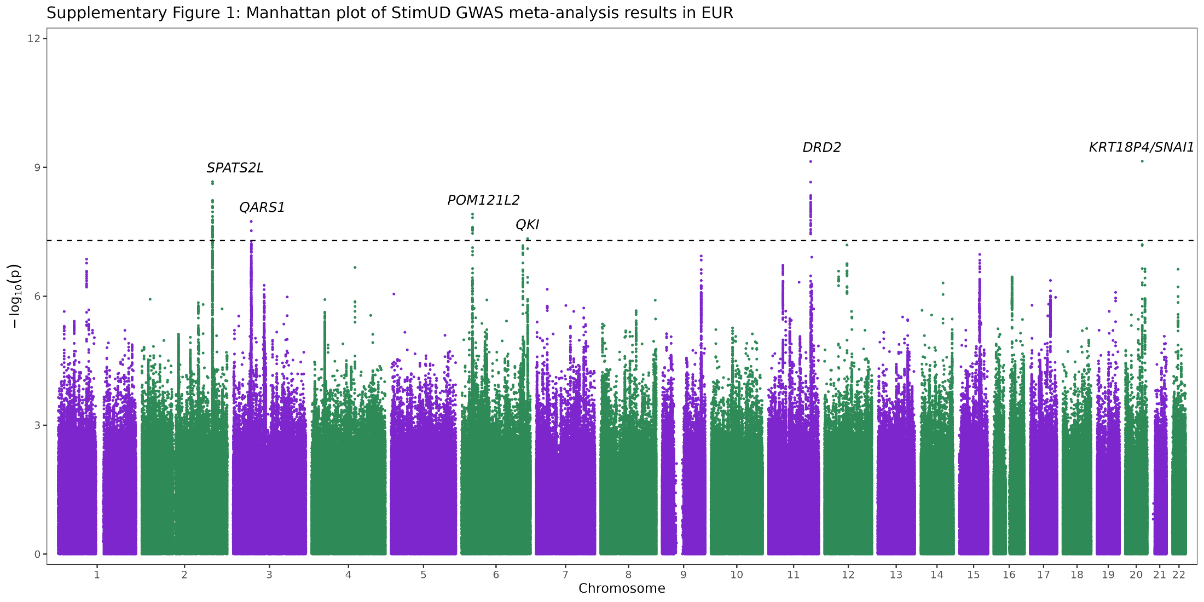

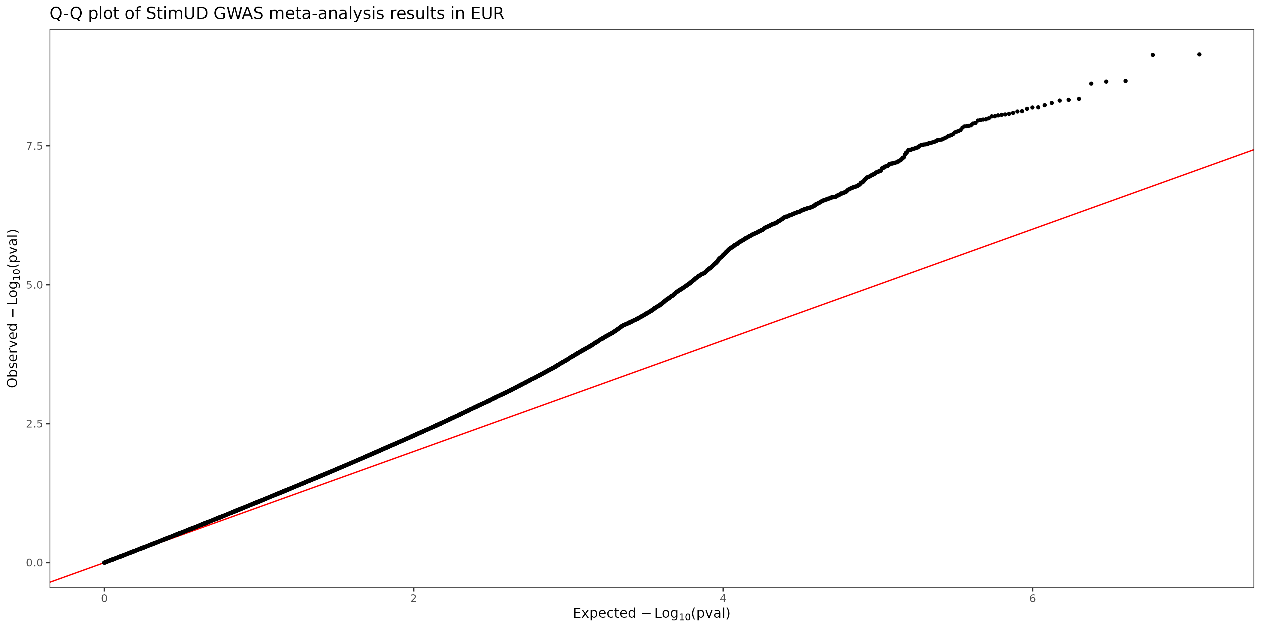

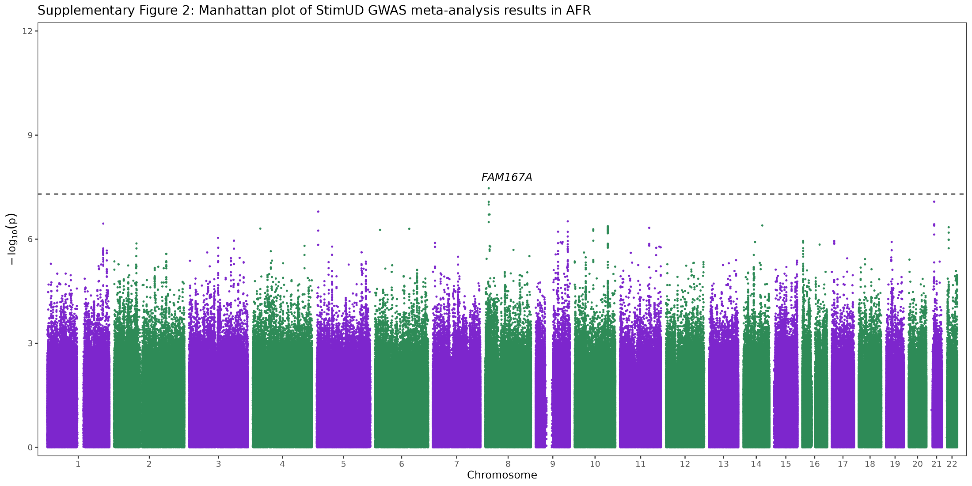

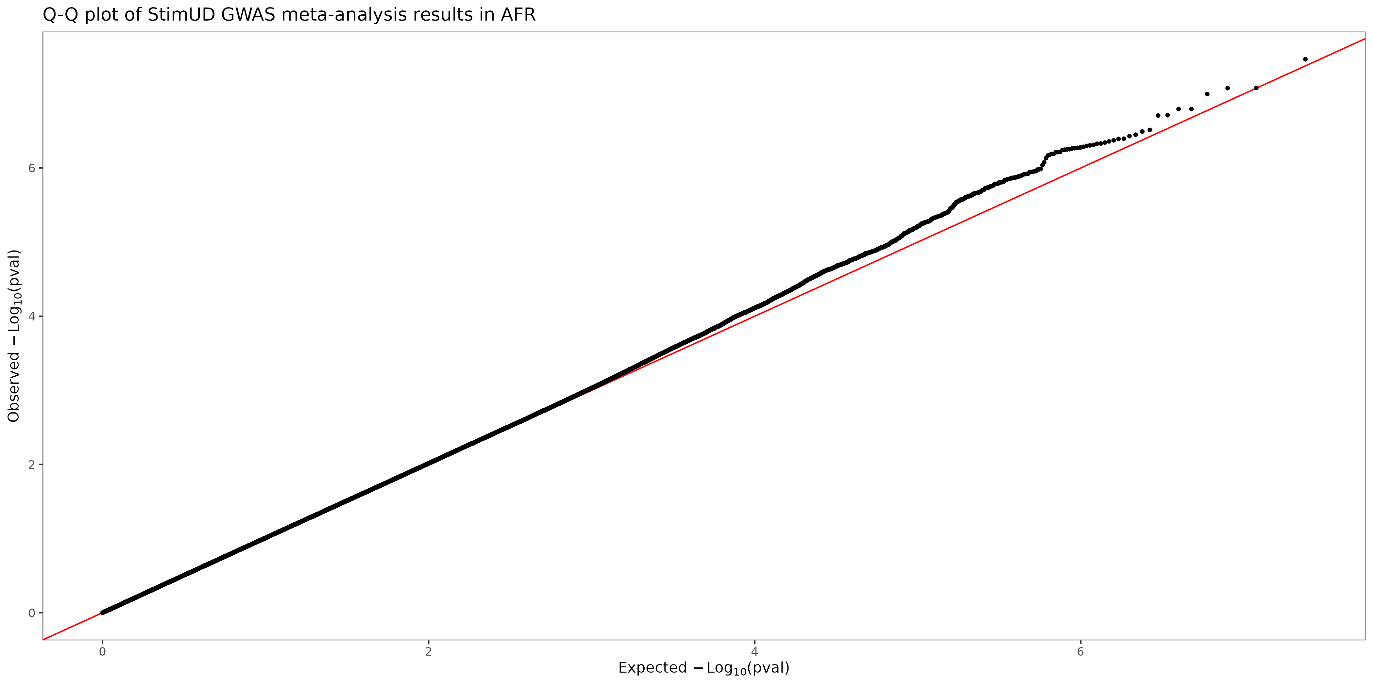

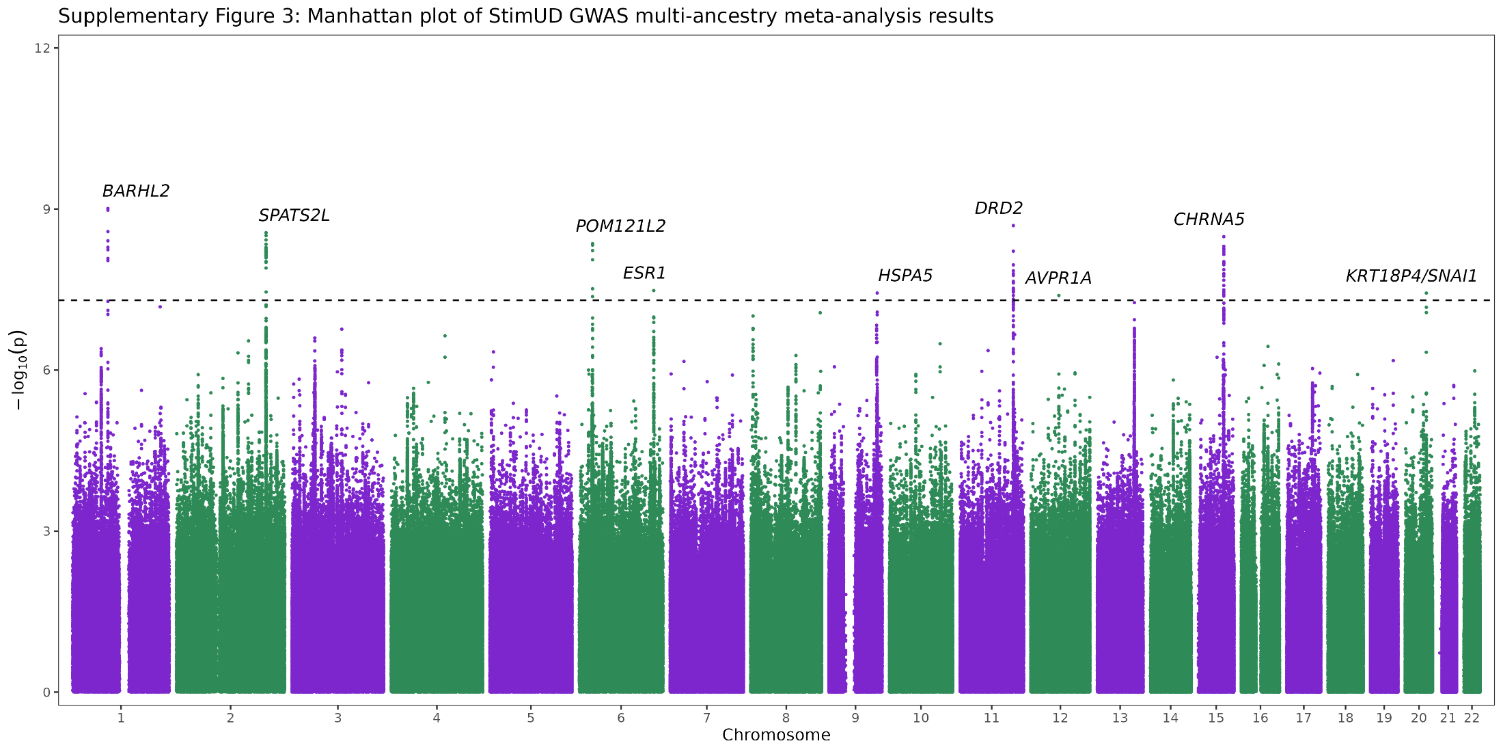

Note: the peak on chromosome 13 approaching significance is *HS6ST3**rs7985691, *P*=5.55×10^-8^.

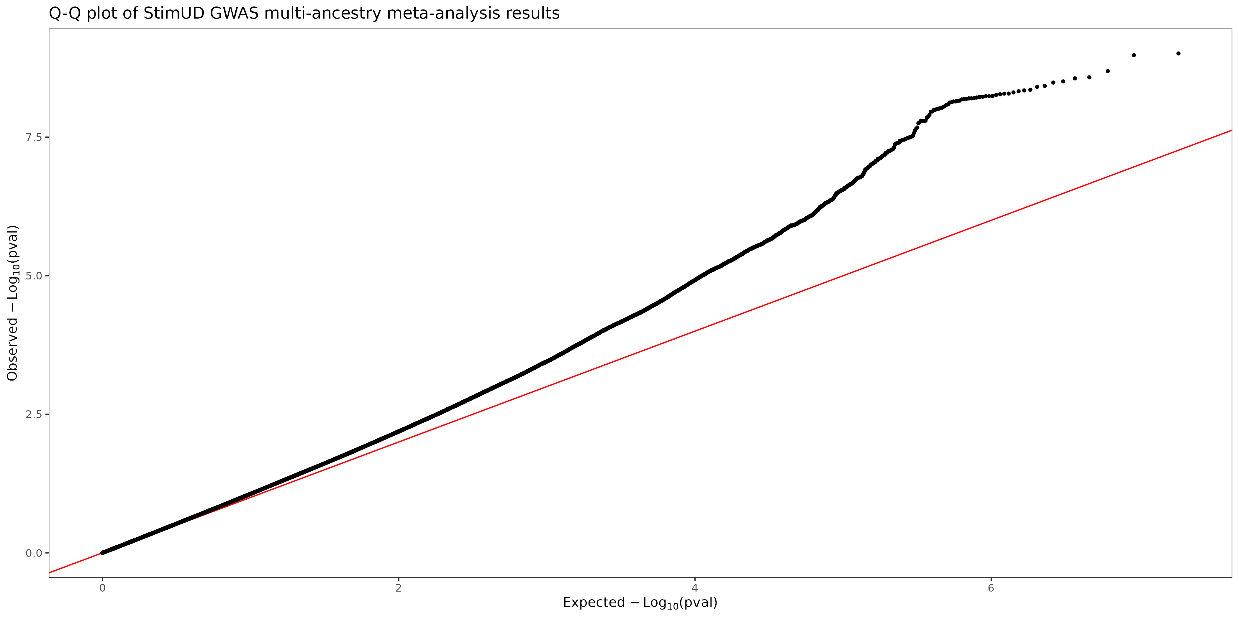

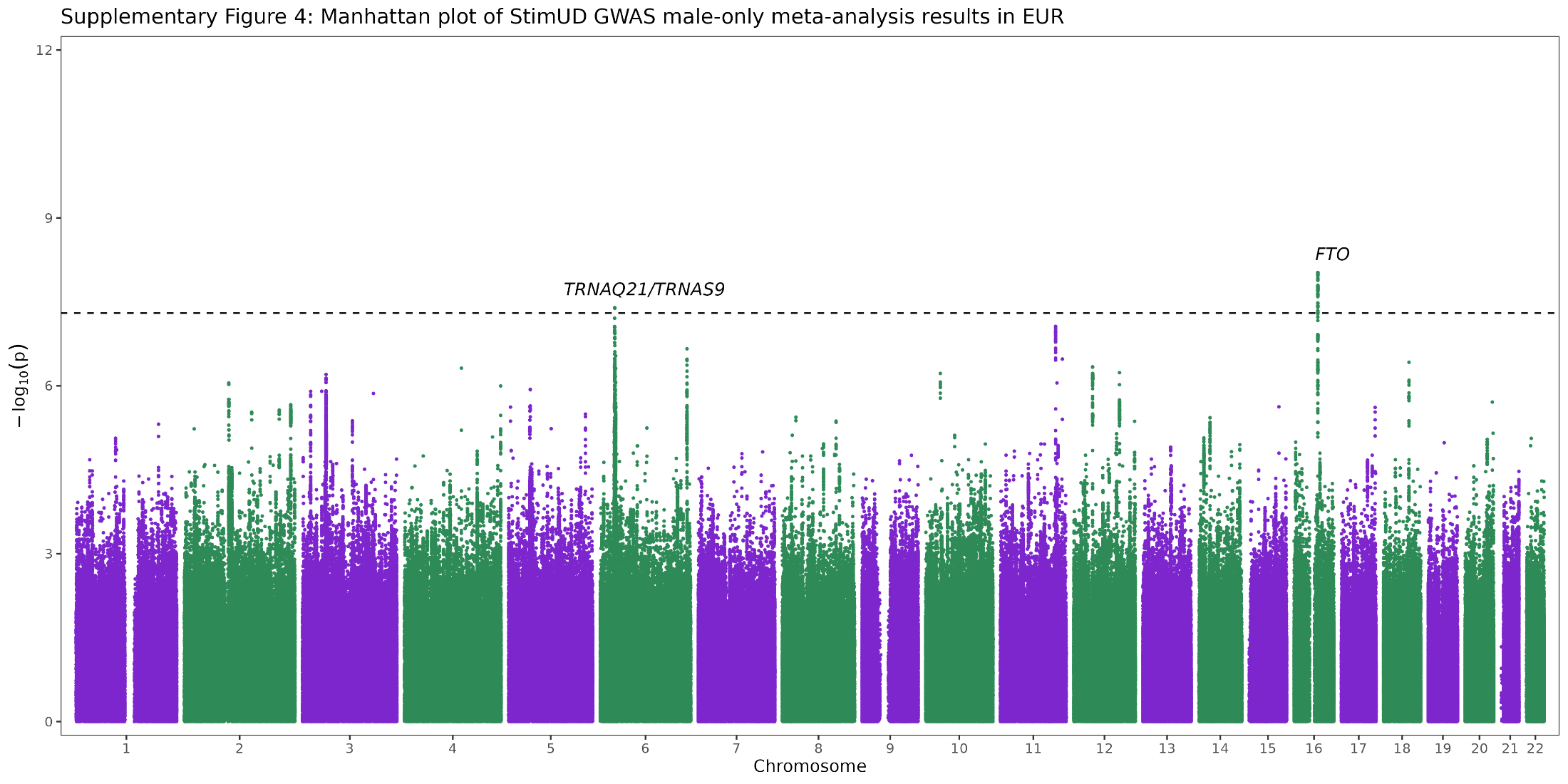

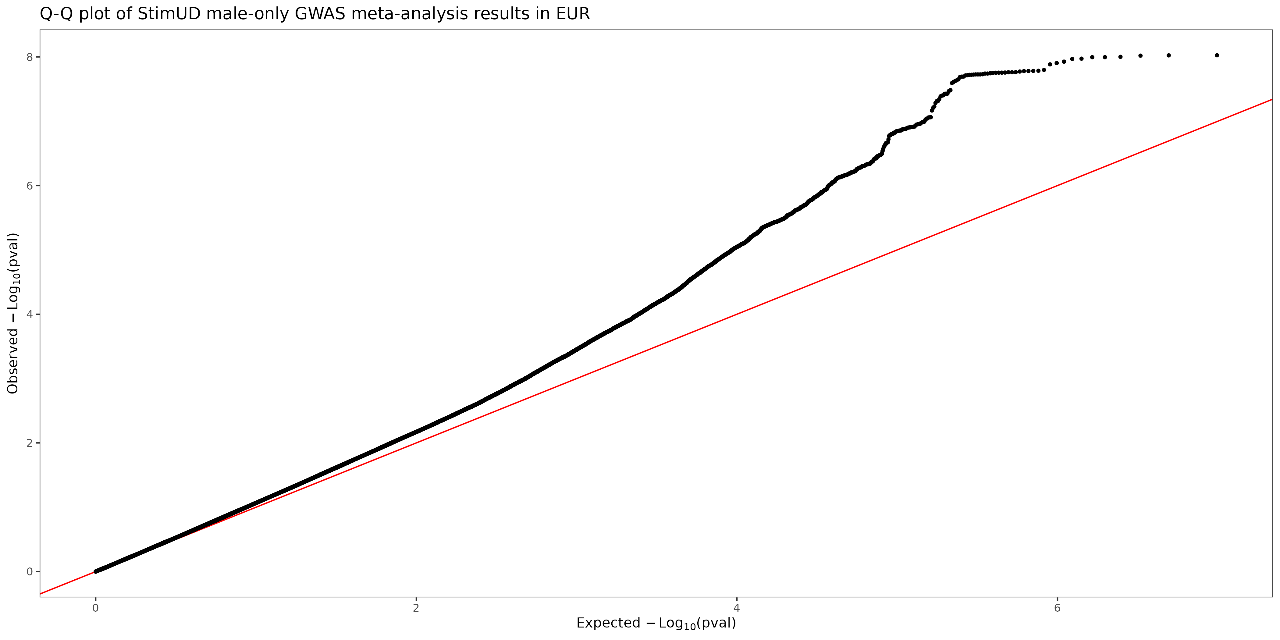

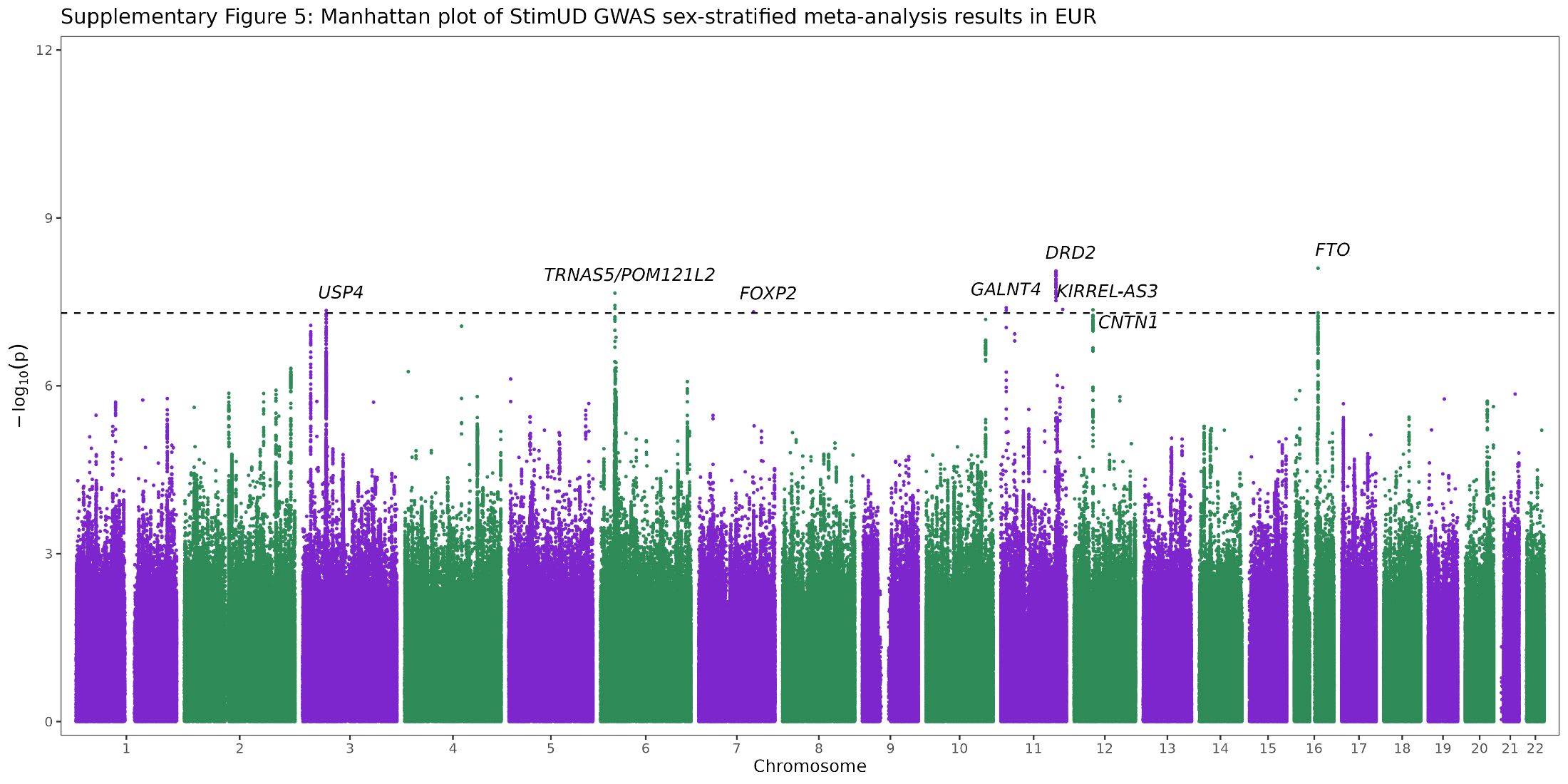

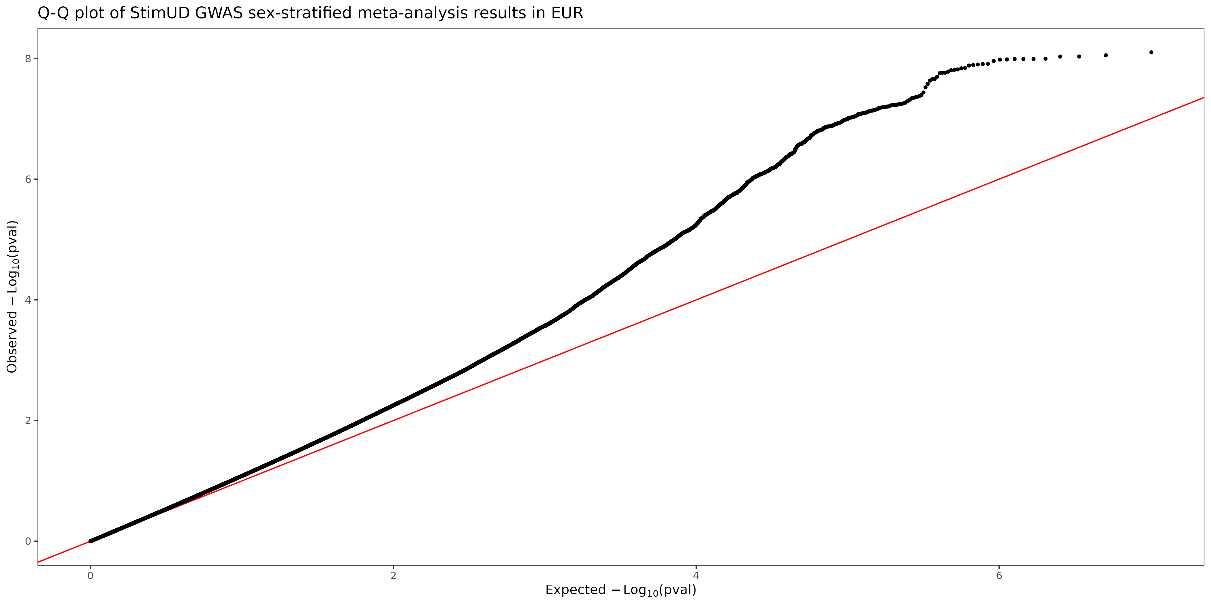

**VA Million Veteran Program Core Acknowledgement
October 2025**

MVP Program Office
Sumitra Muralidhar, Ph.D., Program Director
U.S. Department of Veterans Affairs, 810 Vermont Avenue NW, Washington, DC 20420
Jennifer Moser, Ph.D., Associate Director, Scientific Programs
U.S. Department of Veterans Affairs, 810 Vermont Avenue NW, Washington, DC 20420
Jennifer E. Deen, B.S., Associate Director, Cohort & Public Relations
U.S. Department of Veterans Affairs, 810 Vermont Avenue NW, Washington, DC 20420

MVP Steering Committee
Philip S. Tsao, Ph.D., Co-Chair
VA Palo Alto Health Care System, 3801 Miranda Avenue, Palo Alto, CA 94304
Sumitra Muralidhar, Ph.D., Co-Chair
U.S. Department of Veterans Affairs, 810 Vermont Avenue NW, Washington, DC 20420
J. Michael Gaziano, M.D., M.P.H.
VA Boston Healthcare System, 150 S. Huntington Avenue, Boston, MA 02130
Adriana Hung, M.D., M.P.H.
VA Tennessee Valley Healthcare System, 1310 24th Avenue, South Nashville, TN 37212
Dave Oslin, M.D., Philadelphia VA Medical Center, 3900 Woodland Avenue, Philadelphia,	PA 19104
Deepak Voora, M.D., Durham VA Medical Center, 508 Fulton Street, Durham, NC 27705

MVP Co-Principal Investigators
J. Michael Gaziano, M.D., M.P.H.
VA Boston Healthcare System, 150 S. Huntington Avenue, Boston, MA 02130
Philip S. Tsao, Ph.D.
VA Palo Alto Health Care System, 3801 Miranda Avenue, Palo Alto, CA 94304

MVP Core Operations team
Jessica V. Brewer, M.P.H., Director, MVP Cohort Operations
VA Boston Healthcare System, 150 S. Huntington Ave., Boston, MA 02130
Mary T. Brophy, M.D., M.P.H., Director, VA Central Biorepository
VA Boston Healthcare System, 150 S. Huntington Ave., Boston, MA 02130
Kelly Cho, M.P.H., Ph.D., Director, MVP Phenomics
VA Boston Healthcare System, 150 S. Huntington Ave., Boston, MA 02130
Lori Churby, B.S., Director, MVP Regulatory Affairs
VA Palo Alto Healthcare System, 3801 Miranda Avenue, Palo Alto, CA 94304
Jacob T. Kean, Ph.D., Acting Director, VA Informatics and Computing Infrastructure (VINCI)
VA Salt Lake City Health Care System, 500 Foothill Drive, Salt Lake City, UT 84148
Saiju Pyarajan, Ph.D., Director, Data and Computational Sciences
VA Boston Healthcare System, 150 S. Huntington Avenue, Boston, MA 02130
Robert Ringer, Pharm.D., Director, VA Albuquerque Central Biorepository
New Mexico VA Health Care System, 1501 San Pedro Drive SE, Albuquerque, NM 87108
Luis E. Selva, Ph.D., Director, MVP Biorepository Coordination
VA Boston Healthcare System, 150 S. Huntington Ave., Boston, MA 02130
Shahpoor (Alex) Shayan, M.S., Director, MVP PRE Informatics
VA Boston Healthcare System, 150 S. Huntington Ave., Boston, MA 02130
Brady Stephens, M.S., Principal Investigator, MVP Information Center
Canandaigua VA Medical Center, 400 Fort Hill Avenue, Canandaigua, NY 14424
Stacey B. Whitbourne, Ph.D., Director, MVP Cohort Development and Management
VA Boston Healthcare System, 150 S. Huntington Ave., Boston, MA 02130

15.	Kurki, M.I.J.K., Priit Palta, Timo P. Sipilä, Kati Kristiansson, Kati Donner, Mary P. Reeve, Hannele Laivuori, Mervi Aavikko, Mari A. Kaunisto, Anu Loukola, Elisa Lahtela, Hannele Mattsson, Päivi Laiho, Pietro Della Briotta Parolo, Arto Lehisto, Masahiro Kanai, Nina Mars, Joel Rämö, Tuomo Kiiskinen, Henrike O. Heyne, Kumar Veerapen, Sina Rüeger, Susanna Lemmelä, Wei Zhou, Sanni Ruotsalainen, Kalle Pärn, Tero Hiekkalinna, Sami Koskelainen, Teemu Paajanen, Vincent Llorens, Javier Gracia-Tabuenca, Harri Siirtola, Kadri Reis, Abdelrahman G. Elnahas, Katriina Aalto-Setälä, Kaur Alasoo, Mikko Arvas, Kirsi Auro, Shameek Biswas, Argyro Bizaki-Vallaskangas, Olli Carpen, Chia-Yen Chen, Oluwaseun A. Dada, Zhihao Ding, Margaret G. Ehm, Kari Eklund, Martti Färkkilä, Hilary Finucane, Andrea Ganna, Awaisa Ghazal, Robert R. Graham, Eric Green, Antti Hakanen, Marco Hautalahti, Åsa Hedman, Mikko Hiltunen, Reetta Hinttala, Iiris Hovatta, Xinli Hu, Adriana Huertas-Vazquez, Laura Huilaja, Julie Hunkapiller, Howard Jacob, Jan-Nygaard Jensen, Heikki Joensuu, Sally John, Valtteri Julkunen, Marc Jung, Juhani Junttila, Kai Kaarniranta, Mika Kähönen, Risto M. Kajanne, Lila Kallio, Reetta Kälviäinen, Jaakko Kaprio, Nurlan Kerimov, Johannes Kettunen, Elina Kilpeläinen, Terhi Kilpi, Katherine Klinger, Veli-Matti Kosma, Teijo Kuopio, Venla Kurra, Triin Laisk, Jari Laukkanen, Nathan Lawless, Aoxing Liu, Simonne Longerich, Reedik Mägi, Johanna Mäkelä, Antti Mäkitie, Anders Malarstig, Arto Mannermaa, Joseph Maranville, Athena Matakidou, Tuomo Meretoja, Sahar V. Mozaffari, Mari EK. Niemi, Marianna Niemi, Teemu Niiranen, Christopher J. O’Donnell, Ma’en Obeidat, George Okafo, Hanna M. Ollila, Antti Palomäki, Tuula Palotie, Jukka Partanen, Dirk S. Paul, Margit Pelkonen, Rion K. Pendergrass, Slavé Petrovski, Anne Pitkäranta, Adam Platt, David Pulford, Eero Punkka, Pirkko Pussinen, Neha Raghavan, Fedik Rahimov, Deepak Rajpal, Nicole A. Renaud, Bridget Riley-Gillis, Rodosthenis Rodosthenous, Elmo Saarentaus, Aino Salminen, Eveliina Salminen, Veikko Salomaa, Johanna Schleutker, Raisa Serpi, Huei-yi Shen, Richard Siegel, Kaisa Silander, Sanna Siltanen, Sirpa Soini, Hilkka Soininen, Jae H. Sul, Ioanna Tachmazidou, Kaisa Tasanen, Pentti Tienari, Sanna Toppila-Salmi, Taru Tukiainen, Tiinamaija Tuomi, Joni A. Turunen, Jacob C. Ulirsch, Felix Vaura, Petri Virolainen, Jeffrey Waring, Dawn Waterworth, Robert Yang, Mari Nelis, Anu Reigo, Andres Metspalu, Lili Milani, Tõnu Esko, Caroline Fox, Aki S. Havulinna, Markus Perola, Samuli Ripatti, Anu Jalanko, Tarja Laitinen, Tomi Mäkelä, Robert Plenge, Mark McCarthy, Heiko Runz, Mark J. Daly, Aarno Palotie. FinnGen: Unique genetic insights from combining isolated population and national health register data. (2022).

23.	Bybjerg-Grauholm, J.B.P., Carsten; Bækvad-Hansen, Marie; Giørtz Pedersen, Marianne; Adamsen, Dea; Søholm Hansen, Christine; Agerbo, Esben; Grove, Jakob; Als, Thomas Damm; Schork, Andrew Joseph; Buil, Alfonso; Mors, Ole; Nordentoft, Merete; Werge, Thomas; Børglum, Anders Dupont; Hougaard, David Michael; Mortensen, Preben Bo. The iPSYCH2015 Case-Cohort sample: updated directions for unravelling genetic and environmental architectures of severe mental disorders. (2020).
